# Subjective impact of the COVID-19 pandemic on schizotypy and general mental health in Germany and the UK, for independent samples in May and in October 2020

**DOI:** 10.1101/2021.02.15.21251726

**Authors:** Sarah Daimer, Lorenz Mihatsch, Lisa Ronan, Graham K. Murray, Franziska Knolle

## Abstract

Studies reported a strong impact on mental health during the first wave of the COVID-19 pandemic in March–June, 2020. In this study, we assessed the impact of the pandemic on mental health in general and on schizoptypal traits in two independent general population samples of the UK (May sample N: 239, October sample N: 126; participation at both timepoints: 21) and in two independent general population samples of Germany (May sample N: 543, October sample N: 401; participation at both timepoints: 100) using online surveys. Whereas general psychological symptoms (global symptom index, GSI) and percentage of responders above clinical cut-off for further psychological investigation were higher in the May sample compared to the October sample, schizotypy scores (Schizotypal Personality Questionnaire) were higher in the October sample. We investigated potential associations, using general linear regression models (GLM). For schizotypy scores, we found that loneliness, use of drugs, and financial burden were more strongly corrected with schizotypy in the October compared to the May sample. We identified similar associations for GSI, as for schizotypy scores, in the May and October samples. We furthermore found that living in the UK was related to higher schizotypal scores or GSI. However, individual estimates of the GLM are highly comparable between the two countries. In conclusion, this study shows that while the general psychological impact is lower in the October than the May sample, potentially showing a normative response to an exceptional situation; schizotypy scores are higher at the second timepoint, which may be due to a stronger impact of estimates of loneliness, drug use, and financial burden. The ongoing, exceptional circumstances within this pandemic might increase the risk for developing psychosis in some individuals. The development of general psychological symptoms and schizotypy scores over time requires further attention and investigation.

## 1. Introduction

The highly infectious severe acute respiratory syndrome coronavirus 2 (SARS-CoV-2) had developed into an ongoing worldwide pandemic by March 2020 precipitating a global health crisis with nearly 150 million cases and over 3 million deaths by the end of April 2021 (Daly et al., 2020; JHU, 2021). Due to the high risk of infection and the rapid spread of the virus, governments across the world were compelled to implement restrictions and social distancing measures to keep the number of cases and hospitalisations as low as possible. The main aim of this strategy was to prevent the health care system from being overburdened and gain time to develop treatments and vaccines (Han et al., 2020; Kissler et al., 2020). This led to unpreceded changes to everyday life for the people all around the world. In many countries, people were forced to withdraw from usual face-to-face social activities on a large scale, and schools, nurseries, and retailers as well as workplaces were closed at least for some weeks, with workers being required to work at home. The number of permitted social contacts was limited (Kissler et al., 2020). In many countries, restrictions and lock-down measures are still in place in April 2021. The exact measures taken by countries differed vastly, and even countries within Europe with similar developments throughout the pandemic used different strategies in order to deal with the hitherto unknown situation. According to Plümper and Neumeier (2020) government-strategies can be differentiated based on two dimensions: the time to response and the level of stringency of the lockdown policy. Germany, for example, went into lockdown rapidly in Spring 2020 and managed to control the increase of infections efficiently, whereas the UK delayed lockdown and faced a much higher plateau (Balmford et al., 2020). At the beginning of the pandemic, in March 2020, the government in the UK pondered with the idea of implementing what has since become known as the Swedish strategy, which avoids a lockdown and allows a relatively high number of infections, in order to reach herd immunity (Plümper & Neumayer, 2020). These strategies might have substantially contributed to the variation in numbers of cases and deaths in each country.

From the start of the pandemic, the World Health Organisation, many researchers and clinicians communicated warnings about the consequences of mitigation and suppression measures on mental well-being and mental health (Pfefferbaum and North, 2020; WHO, 2020; Xiong et al., 2020). As expected, the severe restriction of social contacts as well as the fear of the virus or the impact on living conditions had a measurable impact on the mental health of general populations all over the world (Bu et al., 2020; Smith et al., 2020; Xiong et al., 2020). During the first lockdown increased levels of perceived stress and mental distress, COVID-19 related fear, general anxiety and depression and a general decline in mental wellbeing were measured in many countries including, Germany and the UK (Bäuerle et al., 2020; Smith et al., 2020; Proto and Quintana-Domeque, 2021). Female gender, younger age, being part of an ethnic minority and a low socioeconomic status were associated with a high risk for experiencing mental distress (Bäuerle et al., 2020; Fancourt et al., 2020; Simha et al., 2021). However, also living in a specific country was associated with lower stress: for example Adamson and colleagues (2020) found higher perceived stress levels in the UK than in Germany.

The results of our own study from April-May 2020, confirmed these findings revealing a higher psychological and socioeconomic impact of the pandemic on people resident in the UK versus Germany (Knolle et al., 2021). However, both countries reported similarly strong subjective ratings of symptom worsening, with 25% of all responders reported increased levels of anxiety and depressive symptoms (Symptom Check List, SCL-27 (Hardt and Gerbershagen, 2001)), and nearly 10% reported worsening of schizotypy traits measured with Schizotypal Personality Questionnaire (SPQ (Raine, 1991)). Especially, the findings on the subjective worsening of schizotypy measured with the SPQ are interesting, as to our knowledge no other study has investigated schizotypy traits in the general population within the scope of the current pandemic.

Schizotypy describes a latent personality trait, thought to reflect an underlying vulnerability of developing psychosis or schizophrenia-spectrum disorders (Chapman et al., 1994; Debbané and Barrantes-Vidal, 2015; Schultze-Lutter et al., 2019), though we note that different scholars have conceptualised it in differing ways (Grant et al., 2018). Although the SPQ is commonly used as a measure of schizotypy in the general population, its design was based on diagnostic symptoms of schizotypal personality disorder (Oezgen and Grant, 2018). It can, therefore, be considered as measuring related but not identical traits to other schizotypal questionnaires. Based on a recent review by Preti and colleagues (Preti et al., 2020) the current pandemic poses an especially large risk for people suffering from paranoid or high schizotypal traits, as the measures taken to prevent the spread of the virus might ultimately lead to increased anxiety and depressive symptoms, increased avoidance behaviours, stronger disruption of social contacts, and delayed return to normality in these individuals. Furthermore, studies show links between recent adverse life events (Beards et al., 2013; Betz et al., 2020) or isolation and loneliness (Chau et al., 2019; Le et al., 2019) and schizotypy or psychosis-like experiences. Both these aspects, loneliness and adverse life events, are present in the current pandemic, which might have a worsening effect on schizotypal trait expression in people with pre-existing high schizotypy scores, perhaps leading to increased distress or disability. Additionally, the ongoing uncertainty and the impact on social routines might lengthen the time it takes schizotypal high-scores to return to baseline levels. Preliminary evidence and case reports show an increase in the development of first-episode psychosis linked to the pandemic (Huarcaya-Victoria et al., 2020; Valdés-Florido et al., 2020) and reactive psychotic disorders in previously healthy individuals (Valdés-Florido et al., 2020) following the months after start of the pandemic.

When incidence levels of infections decreased during the summer and, as a result, relaxations of the restrictions were initiated (Han et al., 2020; Hetkamp et al., 2020), this also positively impacted the reported mental health status in the general population across both countries. Some studies found a reduction of these scores to a level comparable before the pandemic (Hetkamp et al., 2020; Wang et al., 2020) while others measured elevated, but no longer worsening, levels (Daly et al., 2020; Fancourt et al., 2020; O’Connor et al., 2020). Post-hoc comparisons between countries can be challenging due to the use of different questionnaires by different research groups, or different overall developments of the progression of the pandemic and governmental responses. In this study we therefore, investigated the association of the COVID-19 pandemic and the accompanying lockdown with mental health comparing independent samples of the general population of UK and Germany at two timepoints – the first one during the first lockdown (April/May 2020) and the second after the summer (September/October 2020) when a majority of restrictions had been lifted. Specifically, we examined whether reported levels of depressive symptoms and anxiety, and, in particular, schizotypal scores would change over the summer following the reduction of the restrictions, using the same questionnaire as in the first timepoint. Consistent with other studies, and due to the easing of the restrictions over the summer in Germany and the UK, we hypothesised to detect lower levels of anxiety and depression in the October compared to the May sample. In contrast, we predicted that SPQ-scores would be similar across the first and the second timepoints, as we expected that the return to baseline levels would take longer for schizotypal traits. In addition, all mental health scores were compared between the Germany and the UK to provide insight into the impact of political action on the well-being of the population. For clarity, we wish to emphasize that the design of this study is not longitudinal, rather we assess the impact of the pandemic at two different timepoints in highly comparable but different, only partially overlapping samples.

## 2 Methods

### 2.1. Study design and procedure

The questionnaire used in this study assessing mental and psychological health and COVID-19 exposure was designed as an online survey using EvaSys (https://www.evasys.de, Electric Paper Evaluationssysteme GmbH, Luneburg, Germany). The questionnaire was available in German and English. For participant recruitment we used a snowball sampling strategy to reach the general public. For the first timepoint, data collection took place from 27/04/2020 - 31/05/2020 and completion of the survey took approximately 35 min; for the second timepoint, data collection took place from 10/09/2020 – 18/10/2020, and the completion of the survey took approximately 15 min. For each psychological item, the first timepoint survey included a “before the pandemic” or an evaluation of whether or not item strength had changed, which approximately double the time it took to complete the survey. Participation was voluntary. Participants did not receive any compensation.

Ethical approval was obtained from the Ethical Commission Board of the Technical University Munich (250/20 S). All participants provided informed consent.

### 2.2. Outcome variables

As described in Knolle et al, 2021 in detail, the survey consisted of three parts. The first part, partially comprised of the Coronavirus Health Impact Survey (CRISIS, http://www.crisissurvey.org/), which assessed demographics (age, gender (not biological sex), education and parental education, living conditions), COVID-19 exposure (infection status, symptoms, contact), mental and physical health questions. In the second part, we assessed the general mental health status (global severity of symptom index (GSI)) using the Symptom Check List (SCL) with 27 items (Hardt and Gerbershagen, 2001; Hardt et al., 2011). The GSI score describes the total expression of symptom strength over all SCL-27 items, combining measures of anxiety, depression, mistrust and vegetative symptoms. Furthermore, we assessed the specific sub-scores verified by Hardt and colleagues (2011); the sub-scores were dysthymic symptoms, depressive symptoms, symptoms of social phobia, symptoms of mistrust, agoraphobic symptoms, and vegetative symptoms. In the third part, using the dichotomous version of the Schizotypy Personality Questionnaire (SPQ, (Raine, 1991)) we evaluated total schizotypy trait (SPQ-total). We also assessed the subdimensions, using a six-factor model abnormal experiences and beliefs, social anhedonia, paranoid ideation, social anxiety, eccentricity, and disorganised speech (Davies, 2017), as well as the original nine-factor model (Raine, 1991), the three-factor model (Raine et al., 1994) and the four-factor model (Stefanis et al., 2004). During the first timepoint of data collection, we also assessed subjective measures of change for questions on life circumstances, mental health and psychological traits, asking participants to either report on that particular question before the pandemic or report whether the evaluation of the item had increased, decrease or stayed the same.

### 2.3 Statistical analysis

Statistical analysis and visualisations were computed using R and R Studio (R Core Team, 2016; Rstudio, 2020). We first describe demographics and COVID-19 exposure variables, using frequency analysis. For the country comparison we used Wilcoxon rank sum tests or Chi-square test of independence to explore differences between the countries (UK, Germany) and timepoints (April/ May and September/ October 2020) on the demographics and the COVID-19 exposure variables.

To further explore the differences between the countries and timepoints in CRISIS variables we conducted robust ANOVAS (Mair and Wilcox, 2020) with country (UK, Germany) and timepoint (before pandemic (i.e. subjective rating acquired during the first timepoint), April/ May, September/ October) as between-subjects factor.

In order to identify possible negative associations for experiencing increased general strain and mental distress, we applied Gaussian regression models to assess the connection between the outcome and predictor variables. Our outcome variable was GSI, the total expression of psychological symptoms. In the first basic model, we explored the relationship of demographic variables (age, gender, education, country of residence, living area, parenthood) and prior physical and mental health problems with GSI scores. The second model – the harmful and healthy behaviour model – was used to investigate the link between healthy and harmful behaviours and the outcome variable. For this purpose, we added sleeping hours per night, days with physical exercise per week, drug, alcohol, media consumption and the degree of perceived loneliness to the basic model conducted previously as predictor variables. In the third model – the COVID-19 context model – we examined the coherence of COVID 19 pandemic and associated restrictions using the following variables: perception of the burden of restrictions, stressful relationship changes, financial impact of the Pandemic, hope for a soon end and suspicion of COVID-19 disease. In addition, we also included the degree of trust in government in the October survey.

We investigated possible associations for expression of schizotypy using Poisson regression models. The dependent variable was the total SPQ-score. We used the same three models as for the investigation of the SPQ as for the GSI. We correct for multiple comparisons in all six models, using an adjusted p-value of 0.0083 as the threshold for significance. Models were run for samples separately, using different only partially overlapping samples.

In order to investigate stressful changes in social and family relationships, we used the sum of the degrees of stress experienced in the deterioration in social and family relationships. Excessive media consumption received a positive expression if at least one of the categories of media consumption (television, social media or videogames) was used for more than four hours per day. The drug score was calculated on the basis of at least one use of marijuana, tranquillisers or other drugs like heroin or other opiates.

## 3. Results

### 3.1 Whole Sample Description

The first survey (May 2020) was completed by 860 participants. Two participants did not provide consent and were excluded. 6 participants did not consent to sharing the data publicly, and are included in the analysis but will be removed from the open-access data set. In this paper we focused on the comparison of responders living in the UK (N=239) and in Germany (N=543). In the first survey the majority of respondents were female (72%), 25% were male, 3% diverse or did not provide the information. The age ranged from 18 to 92 years, with a mean 43 (SD 15.5) years and a median of 41 years. The majority of participants were well educated, 60% had a master’s degree or higher, and 25% had completed a professional college or a bachelor degree. 48% reported to live in large cities, 12% in suburbs of large cities, 19% in small cities, and 21% in rural areas. (See Table 1).

**Table 1.**
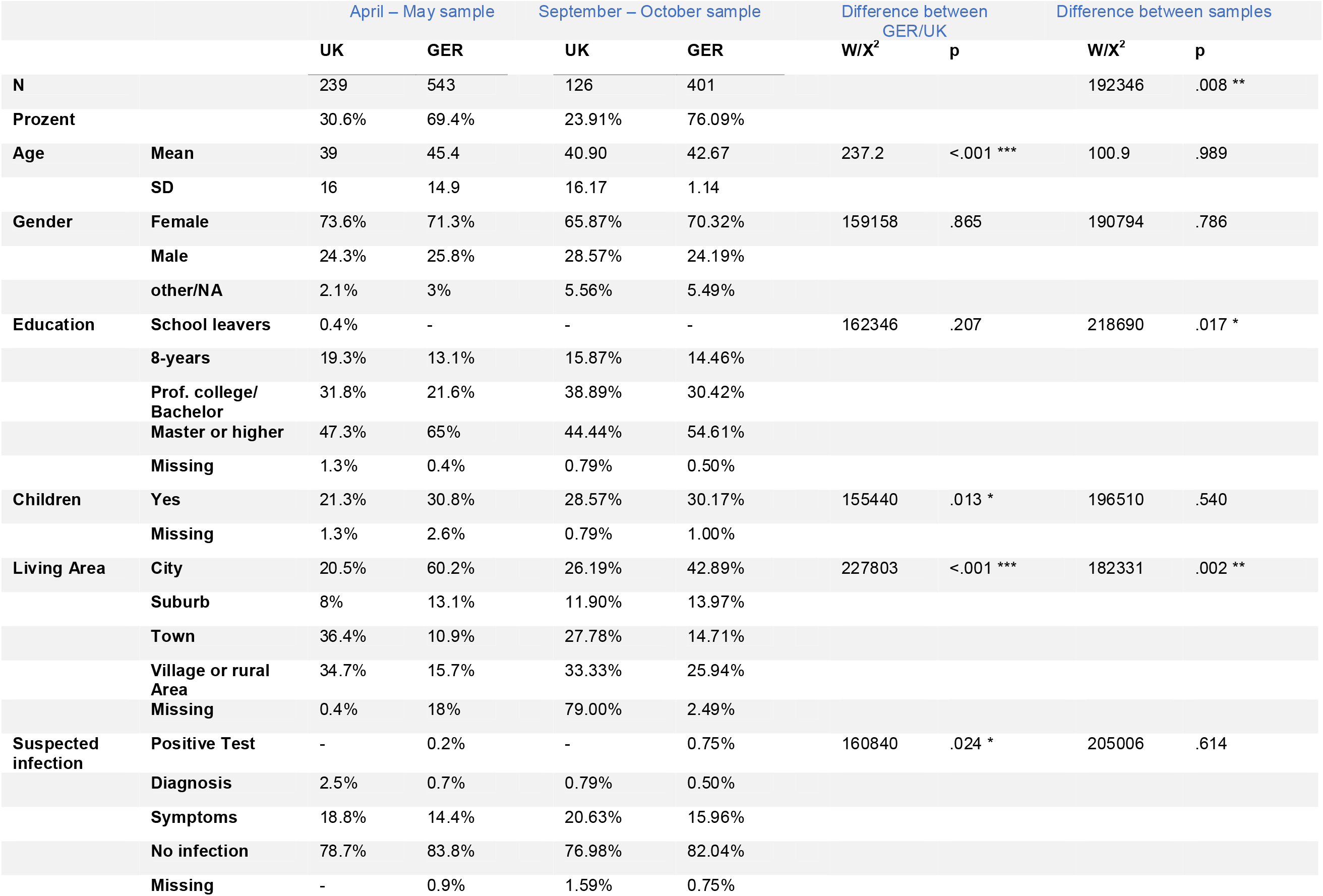
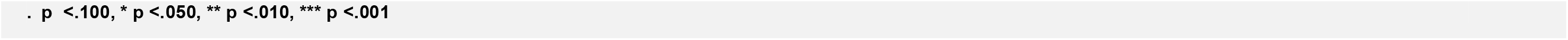
Demographics and suspected infection among the samples divides by country.

|  |  | April – May sample |  | September – October sample |  | Difference between<br>GER/UK |  | Difference between samples |  |
| --- | --- | --- | --- | --- | --- | --- | --- | --- | --- |
|  |  | UK | GER | UK | GER | W/X<sup>2</sup> | p | W/X<sup>2</sup> | p |
| **N** |  | 239 | 543 | 126 | 401 |  |  | 192346 | .008 ** |
| **Prozent** |  | 30.6% | 69.4% | 23.91% | 76.09% |  |  |  |  |
| **Age** | **Mean** | 39 | 45.4 | 40.90 | 42.67 | 237.2 | <.001 *** | 100.9 | .989 |
|  | **SD** | 16 | 14.9 | 16.17 | 1.14 |  |  |  |  |
| **Gender** | **Female** | 73.6% | 71.3% | 65.87% | 70.32% | 159158 | .865 | 190794 | .786 |
|  | **Male** | 24.3% | 25.8% | 28.57% | 24.19% |  |  |  |  |
|  | **other/NA** | 2.1% | 3% | 5.56% | 5.49% |  |  |  |  |
| **Education** | **School leavers** | 0.4% | - | - | - | 162346 | .207 | 218690 | .017 * |
|  | **8-years** | 19.3% | 13.1% | 15.87% | 14.46% |  |  |  |  |
|  | **Prof. college/**<br>**Bachelor** | 31.8% | 21.6% | 38.89% | 30.42% |  |  |  |  |
|  | **Master or higher** | 47.3% | 65% | 44.44% | 54.61% |  |  |  |  |
|  | **Missing** | 1.3% | 0.4% | 0.79% | 0.50% |  |  |  |  |
| **Children** | **Yes** | 21.3% | 30.8% | 28.57% | 30.17% | 155440 | .013 * | 196510 | .540 |
|  | **Missing** | 1.3% | 2.6% | 0.79% | 1.00% |  |  |  |  |
| **Living Area** | **City** | 20.5% | 60.2% | 26.19% | 42.89% | 227803 | <.001 *** | 182331 | .002 ** |
|  | **Suburb** | 8% | 13.1% | 11.90% | 13.97% |  |  |  |  |
|  | **Town** | 36.4% | 10.9% | 27.78% | 14.71% |  |  |  |  |
|  | **Village or rural**<br>**Area** | 34.7% | 15.7% | 33.33% | 25.94% |  |  |  |  |
|  | **Missing** | 0.4% | 18% | 79.00% | 2.49% |  |  |  |  |
| **Suspected**<br>**infection** | **Positive Test** | - | 0.2% | - | 0.75% | 160840 | .024 * | 205006 | .614 |
|  | **Diagnosis** | 2.5% | 0.7% | 0.79% | 0.50% |  |  |  |  |
|  | **Symptoms** | 18.8% | 14.4% | 20.63% | 15.96% |  |  |  |  |
|  | **No infection** | 78.7% | 83.8% | 76.98% | 82.04% |  |  |  |  |
|  | **Missing** | - | 0.9% | 1.59% | 0.75% |  |  |  |  |

. $p < .100$ , \* $p < .050$ , \*\* $p < .010$ , \*\*\* $p < .001$

The second survey (October 2020) was completed by 550 participants. 22 were excluded from the analysis as they gave not consent to the participation and one he or she did not provide information about the current residency. 69 % of the participants were female, 25% male and 6% did not provide the information. The age ranged from 18 to 93 years (M= 42, SD=16.1). Most of the participants had their current residency in Germany (76%, N=401) and 24% in the UK (N=126). The majority of the sample were well educated, with 45% reporting to have a master’s degree or higher and 39% to have completed a professional college or bachelor’s degree. 39% lived in a city, 13% in suburbs, 18% in towns and 28% in villages or rural areas. (See Table 1).

Since participation at the first survey was not required for taking part in the second survey, the two samples are partially overlapping. 121 responders participated in both surveys. The samples did not differ significantly in terms of age (X^2^=100.8, p=.989) and gender (W = 192786, p = .635) between the timepoints, but in the second survey significantly more participants came from Germany (X^2^= 8.55, p = .014), their living area was more rural (W = 182331, p=.002) and the sample was less educated (W = 218690, p = .018).

### 3.2 COVID 19 infection

At the first timepoint, 0.2% of the German sample reported a positive COVID-19 test result, 0.7% reported a diagnosis made by a health care professional without using a test, but based on symptoms and contact to COVID-19 positive individuals, and 14.4% possible symptoms of a COVID-19 infection. 83,8% stated that they had not suspected COVID-19. In the UK sample, 2.5% reported a medical diagnosis of COVID-19 made by a health care professional without using a test, but based on symptoms and contact to COVID-19 positive individuals, and 18.8% of possible symptoms. None of the respondents reported having received a positive test result. 78.7% reported that they had not previously suspected COVID-19.

At the second timepoint, 0.8% of the German sample reported being positively tested for COVID-19, 0.5% reported receiving a positive diagnosis, without a test, and 16% reported symptoms that could indicate a COVID-19 infection. 82.0% reported not having suspected COVID-19. In the UK sample, 0.8% reported having received a positive diagnosis, without a test, of COVID-19 and 20.6% had recently experienced symptoms of COVID-19 infection. None of the UK respondents reported having received a positive test result. 77.0% reported no signs of possible COVID-19 infection. (See Table 1).

### 3.2 Results of robust ANOVAs

GSI scores differed significantly between the two countries (p <.001) and samples (p =.04) with higher GSI scores in the May sample and higher scores in the UK sample compared to the German sample (see Figure 1). There was no interaction effect. The results of the robust ANOVAS are shown in Table 2. Furthermore, we investigated the development in the small sample of those responders who took part in the survey at both timepoints. The results are similar and are presented in the supplementary materials.

**Table. 2.**
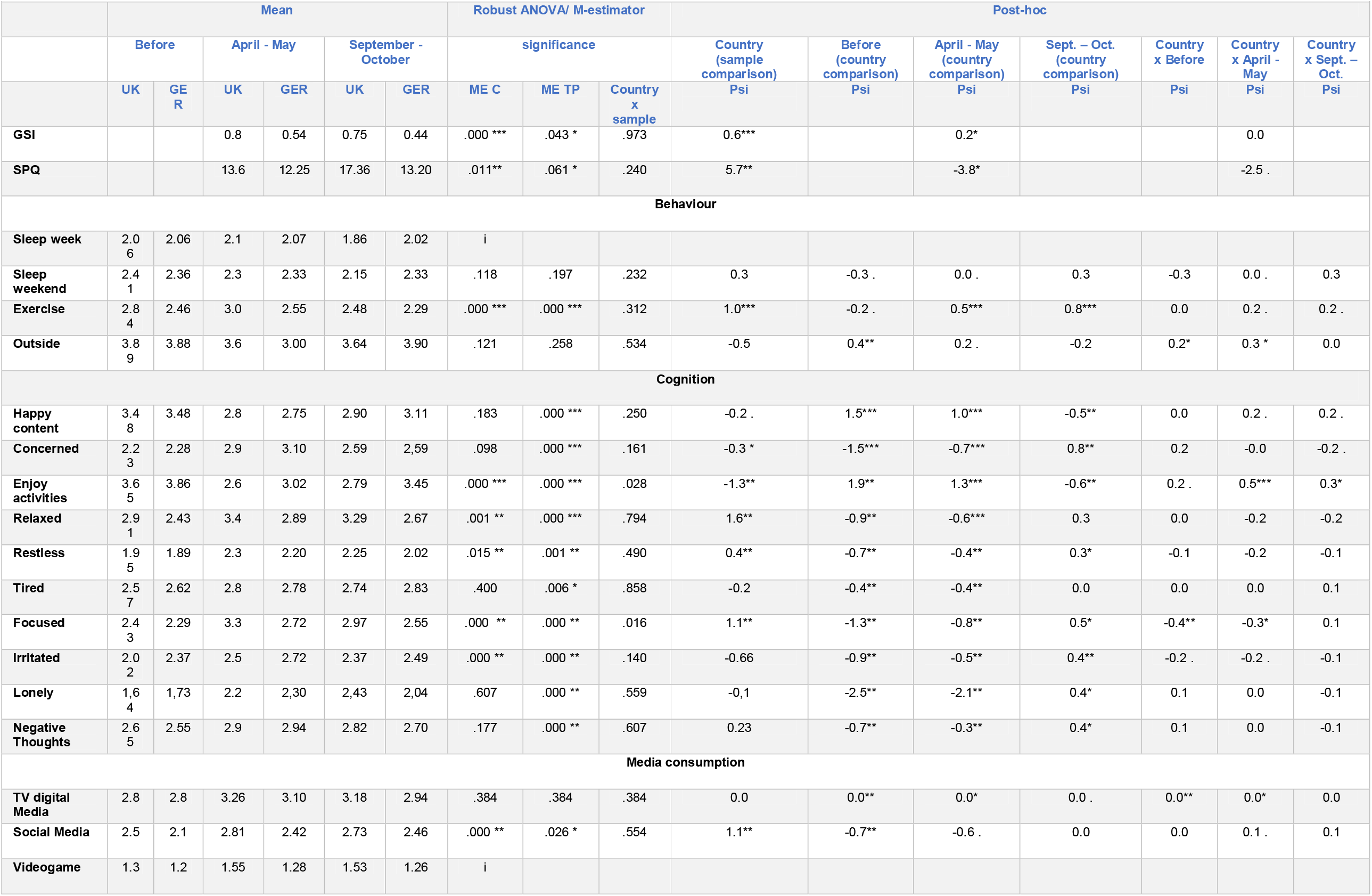
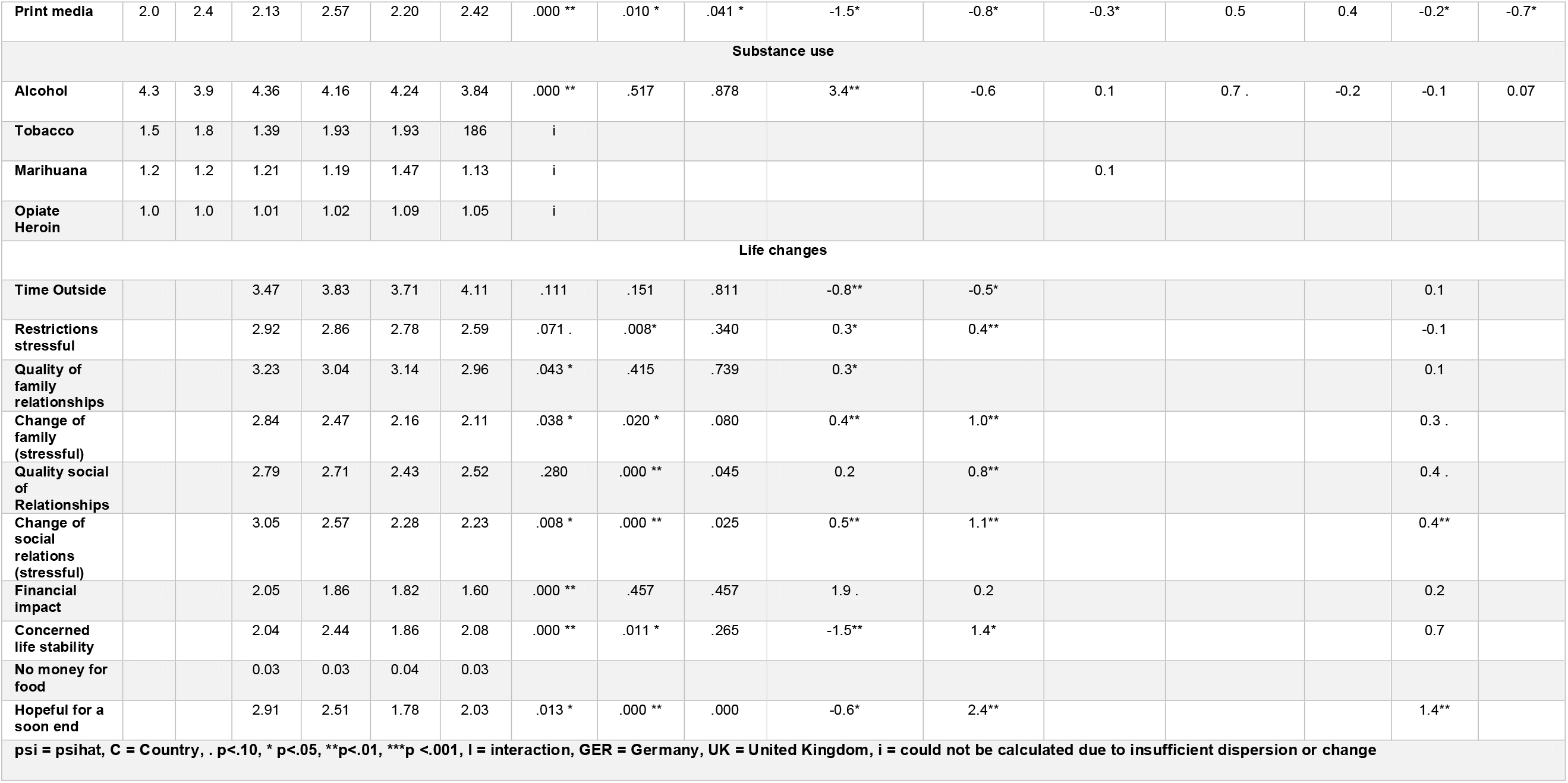
Overview of means and robust ANOVAS of GSI and SPQ scores, all CRISIS variables and questions concerning life changes due to COVID.

**Figure 1:**
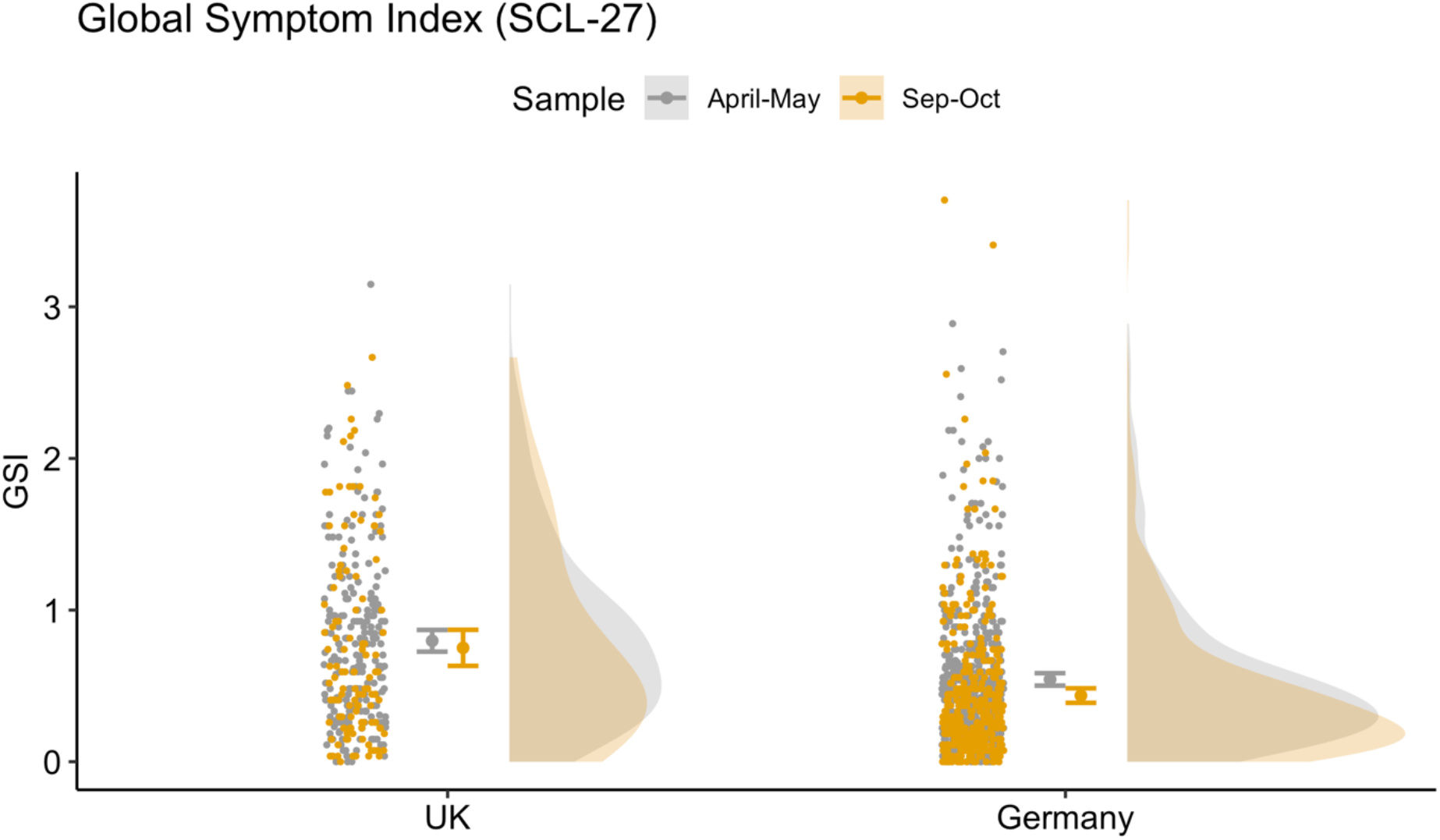
Raincloud plot for GSI across country and timepoint, showing data distribution (the ‘cloud’), jittered raw data (the ‘rain’), mean and standard error.

In a norm sample (Hardt et al., 2004), 10-15% of the screened population reach the clinical cut-off on the different sub-dimensions, and require additional psychological investigation. As shown in Figure 2, there are significant differences between the countries (F(1, 7464 = 237.96, p <.001) and samples (F(1, 7464 = 12.58, p <.001). For the sub-dimension of dysthymic symptoms (DYS), the rate fell from 68.38% to 58.82% in the UK responders and from 37% to 30% in the German responders who lay above the clinical cut-off; for depressive symptoms (DEP) from 51% to 50% in the UK and from 39% to 27% in the German responders; for symptoms of social phobia (SOP) from 37% to 34% in the UK and from 24% to 19% in German responders; for symptoms of mistrust (MIS) from 29% to 26% in the UK and from 26% to 22% in the German sample; and for agoraphobic symptoms (AGO) from 52% to 32% in the UK and from 23% to 12% in the German responders. Interestingly, the vegetative symptoms (VEG) increased from 26% to 35% in the UK and from 14% to 16% in the German responders. The reduced rates were measured in a comparable but different only partially overlapping sample.

**Figure 2:**
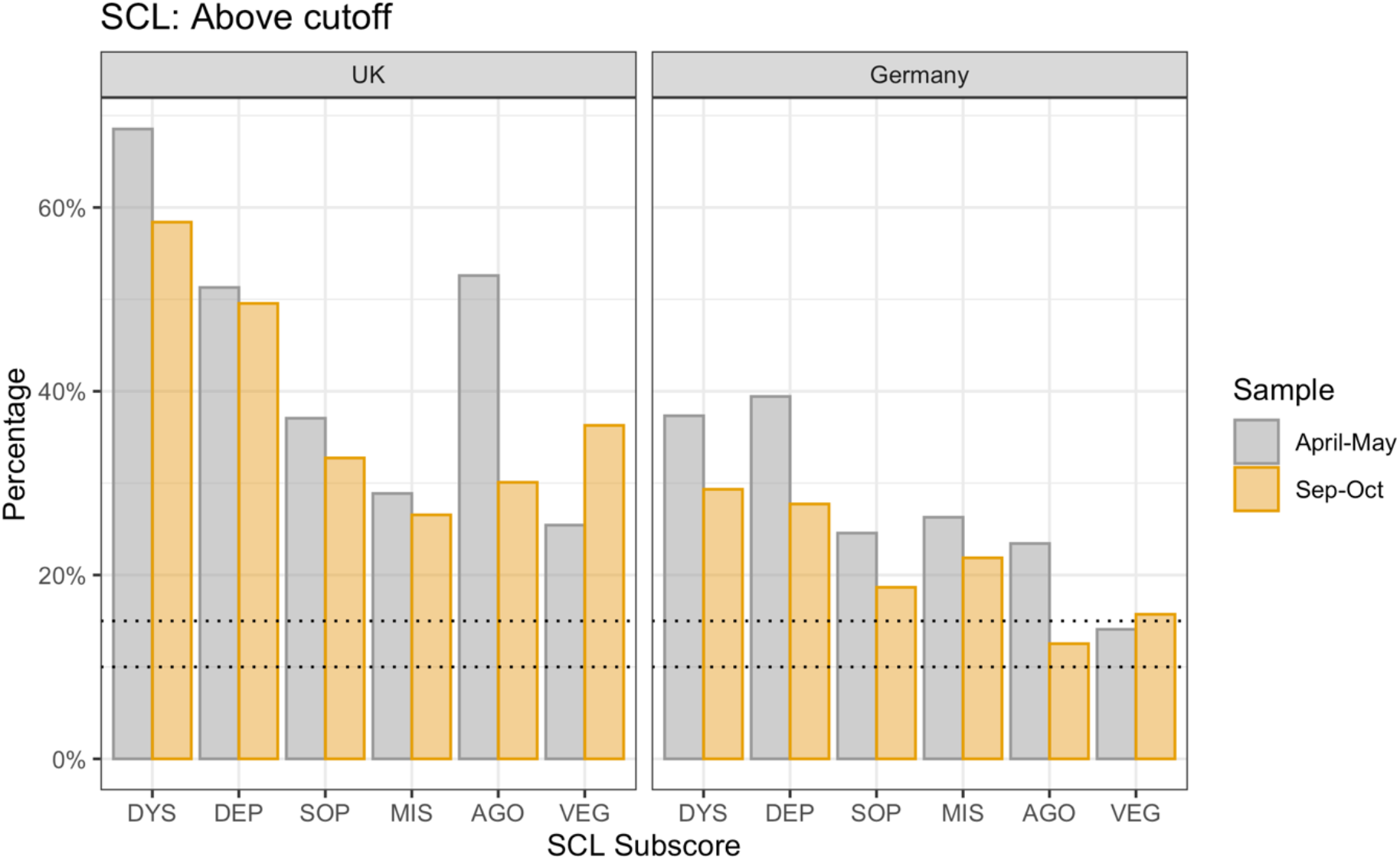
Percentage of responders above clinical cut-off by country and sample. Dotted lines represent the percentage of the norm population above threshold (10-15%). DYS: dysthymic symptoms, DEP: depressive symptoms, SOP: symptoms of social phobia, MIS: symptoms of mistrust, AGO: agoraphobic symptoms, VEG: vegetative symptoms.

SPQ scores (Figure 3) also differed significantly between countries with higher scores in the UK sample (p =.01). We found a trend towards higher SPQ-scores in the October compared to the May sample (p = .06). The results of the robust ANOVAS are shown in Table 2. Furthermore, we investigated the development of the total SPQ score in the small sample of those responders who took part in the survey at both timepoints. The results are similar and are presented in the supplementary materials.

**Figure 3:**
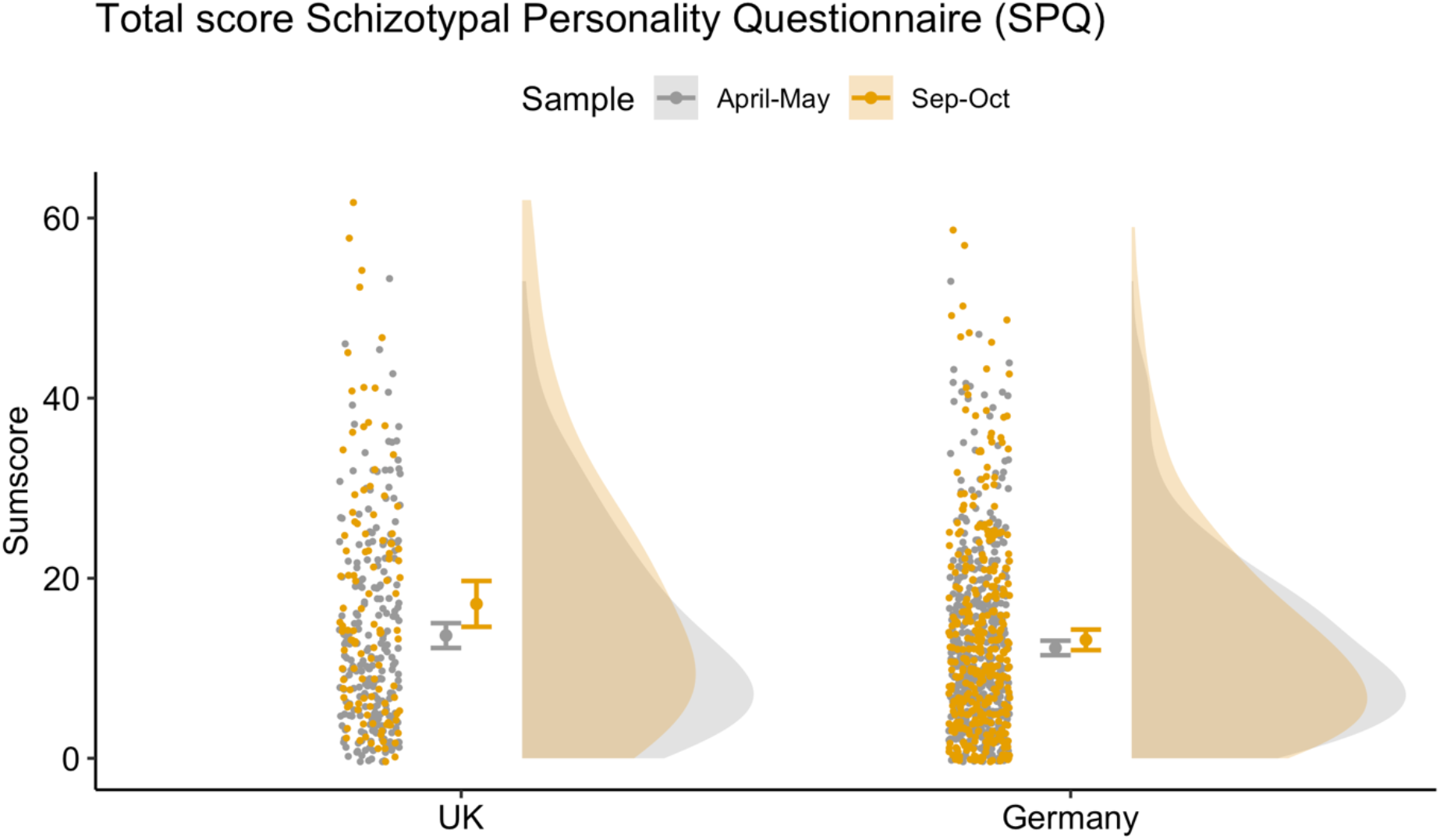
Raincloud plot for total SPQ across country and sample, showing data distribution (the ‘cloud’), jittered raw data (the ‘rain’), mean and standard error.

Additionally, we investigated the different SPQ subdimensions in an explorative analysis. For completion, we present four different categorisations based on the six-factor model ((Davies, 2017) Figure 4A), the original nine factor model ((Raine, 1991), Figure 4B), the three-factor model ((Raine et al., 1994), Figure 4C) and the four-factor model ((Stefanis et al., 2004), Figure 4D). Using a robust ANOVA, we analysed differences across country of residence and samples. All results are presented in the supplementary Table 1. Factors of social anhedonia and social anxiety (Raine 1991, Davies 2017), as well as the interpersonal factor (Stefanis et al., 2004; Raine et al., 1994) show the strongest endorsement in the two countries. The overall trend is similar across all four approaches, revealing significant differences between the two countries and showing higher scores for most subdimension scores in the UK.

**Figure 4:**
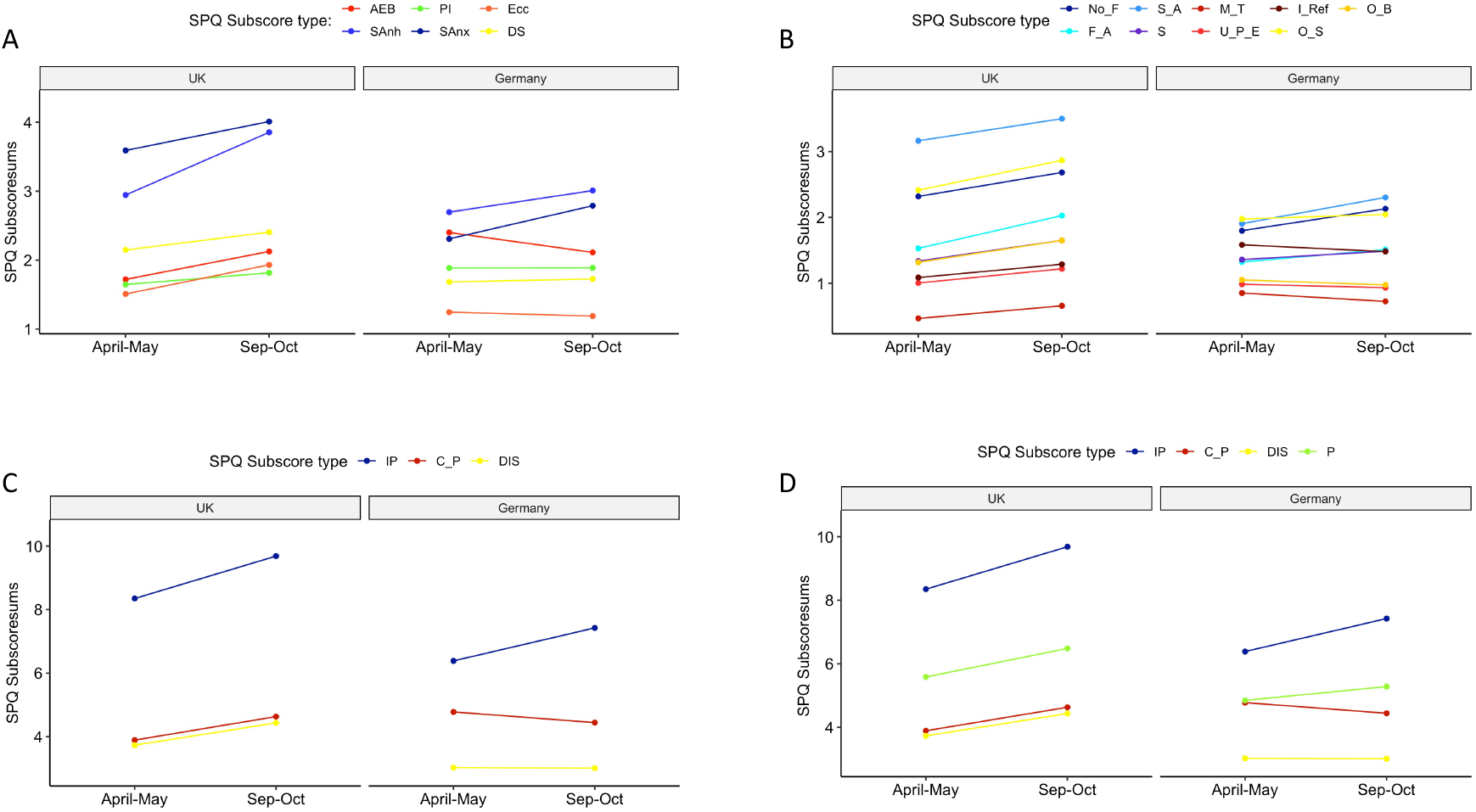
Interaction plot shows different subdimension models across samples and countries. A. Six-factor model by Davies et al. (2017). AEB: Anomalous experiences and beliefs, SAnh: Social anhedonia, PI: Paranoid ideation, SAnx: Social anxiety, Ecc: Eccentricity, DS: Disorganised speech. **B. Nine-factor model by Raine, 1991**. No_F: No close friends, F_A: Flattened/ Constricted affect, S_A: Social anxiety, S: Suspiciousness, M_T: Magical thinking, U_P_E: Unusual perceptual experience, I_Ref: Ideas of Reference, O_S: Odd speech, O_B: Odd behavior. **C. Three-factor model by Raine et al. (1994)**. IP: Interpersonal, C_P: Cognitive/ Perceptual, DIS: Disorganization. **D. Four-factor model by Stefanis et al. (2004)** with additional Paranoid (P) subdimension.

### 3.3 General linear model: General psychological symptom index

#### 3.3.1 Effects of Demographic Variables and prior physical and mental health problems on GSI scores (Basic Model)

In both surveys in May and October, age (TP1: p <.001, TP2: p = .001), country of residence (TP1 p = .005, TP2 p <.001) and pre-existing physical (TP1: p =.002, TP2: p = 0.005) and mental health problems (TP1: p <.001, TP2: p <.001) were significantly associated with GSI. Older age and country of residence in Germany are related to lower scores, while the opposite was shown for pre-existing physical and mental health problems. Female gender was associated with lower GSI scores in the May sample (TP1: p =.031, TP2: p = .779) and higher levels of education was related to lower scores in the October sample (TP1: p =.962, p <.001). In the first survey we found that living in a town was significantly connected to higher GSI compared to living in a large city (p =.045). There was no significant association between having children at home and the outcome in both samples in May and October (TP1: p =.256, TP2: p =.439). See Table 3.

**Tab. 3.**
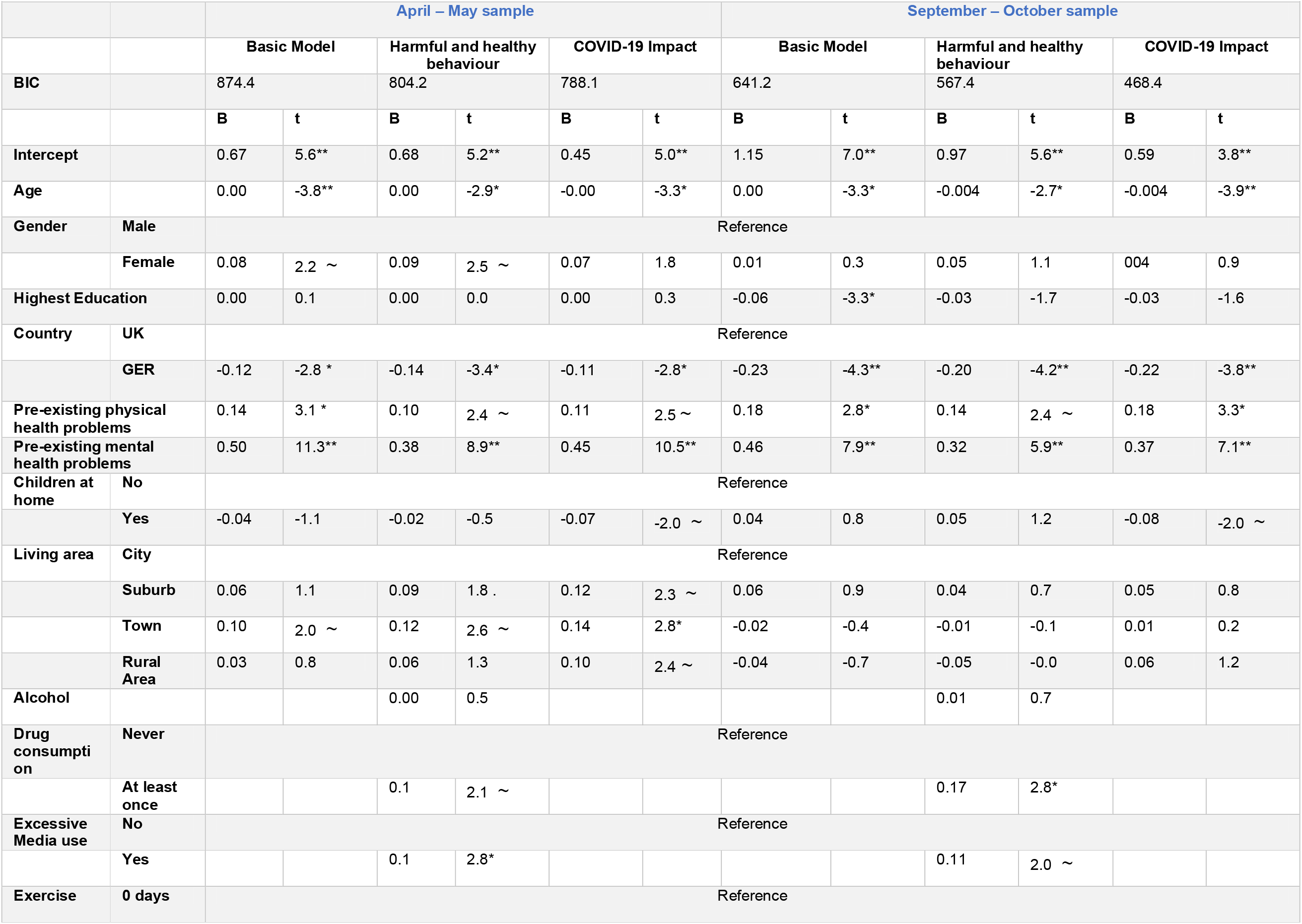
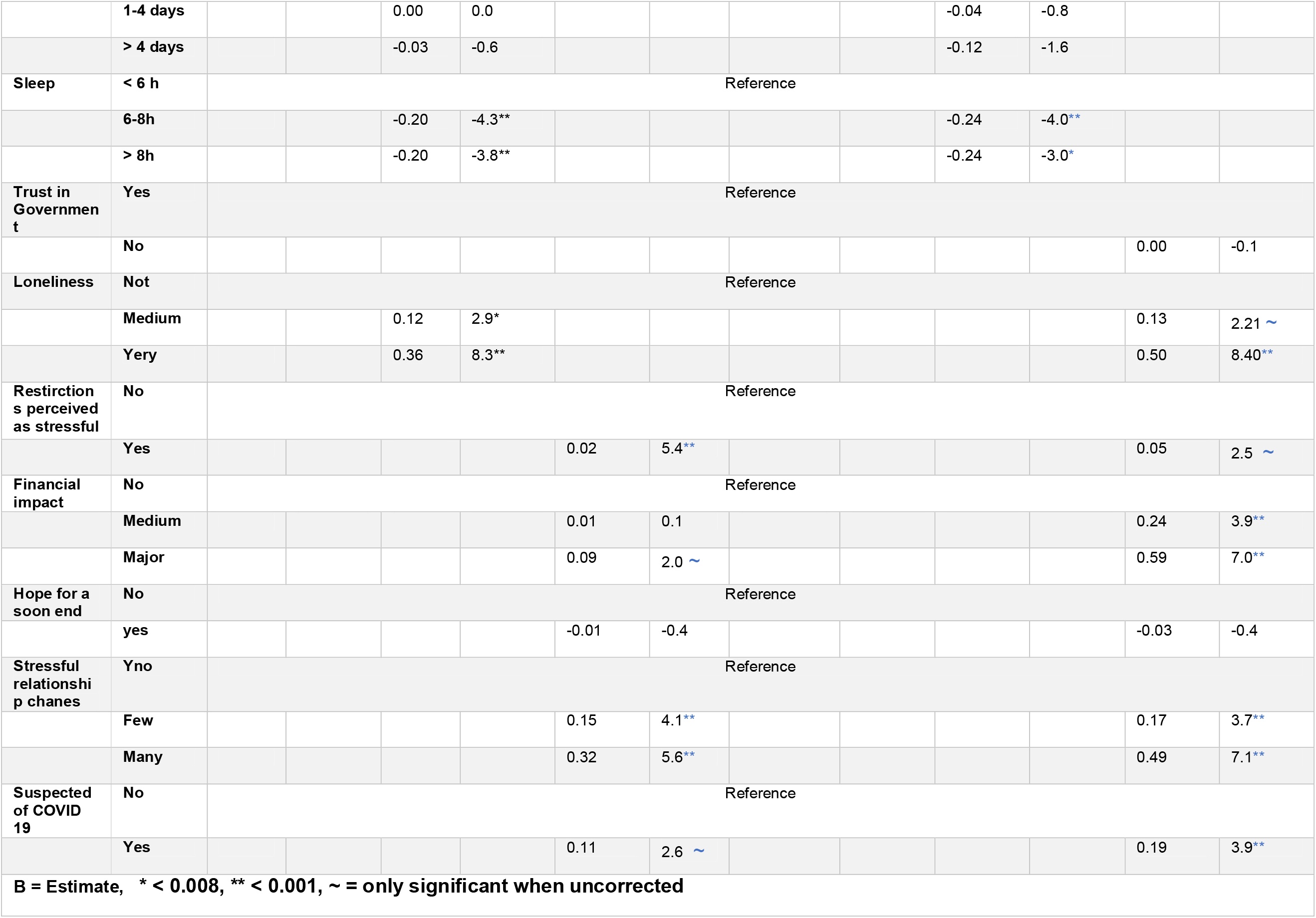
Overview over all three conducted models (Basic Model, Harmful and Healthy Behavior, COVID-19 Impact) for both samples for GSI scores.

Based on these results, we adjusted for the predictors age, gender, education level, country of residence, having children at home, as well as physical and mental health problems in the following two models. Additionally, all models have been run separately on the small overlapping sample, see supplementary materials. Results are comparable to the full sample.

#### 3.3.2 Effects of harmful and healthy behaviours GSI scores (Harmful and Healthy Behaviour Model)

After adjusting for possibly confounding demographic variables from the previous basic model, in the May and October samples drug consumption was associated with higher GSI scores (TP1: p = .005, TP2: p <.001), as well as excessive media use (TP1: B = 0.12, t = 3.34, p <.001, TP2: B =0.13, t = 2.38, p <.001). No correlation was found for alcohol consumption (TP1: p = .602, TP2: p =.740). Sleeping between 6 and 8 hours (TP1: p <.001, TP2: p <.001) and more than 8 hours (TP1: p<.001, TP2: p <.001) compared to sleeping less than 6 hours was connected to lower GSI scores. There was no effect of physical exercise on GSI. Feeling lonely had a negative association with GSI scores both on medium levels (TP1: p =.046, TP2: p <.001) and high levels (TP1: p =.003, TP2: p <.001) in the May and October samples. See Table 3 and supplementary materials for results of the fully overlapping sample.

#### 3.3.3 Effects of the COVID-19 pandemic on GSI scores (COVID-19 Impact Model)

In this third model, we investigated the relationship of factors related to the COVID-19 pandemic with GSI scores. The perception of the restrictions as being stressful was connected with increased GSI scores in the May and October samples (TP1: p <.001, TP2: p =.013). Financial problems due to the crisis significantly correlated with GSI scores, being higher in the May compared to the October survey. In the May samples, only major financial impact was associated with increased GSI scores (p =.042). In the October samples, both major financial impact (p <.001) and medium impact (p <.001) were negatively related to GSI. Deteriorations in relationships that were experienced as stressful had a negative connection with the outcome in both, the May and October samples, with very stressful changes having a greater association (TP1: p <.001, TP2: p <.001) on GSI than stressful changes only (TP1: p <.001, TP2: p<.001). The suspicion of COVID-19 disease or the diagnosis had a negative relationship with GSI scores in both, the May and October samples (TP1: p =.009, TP2: p <.001). Being hopeful for a soon end of the pandemic did not have a significant association with GSI scores. In the October sample, we also included the degree of trust in the government to lead the country well out of the crisis in our model. However, this predictor had no significant effect on GSI scores. See Table 3 and supplementary materials for results of the fully overlapping sample.

### 3.4 General linear model: Schizotypy (SPQ)

#### 3.4.1 Effects of Demographic Variables and prior physical and mental health problems on SPQ scores (Basic Model)

In both, the May and October samples, increasing age (TP1p <.001, TP2: <.001), higher education levels (TP1: p <.001, TP2: p<.001) and female gender (TP1: p =.088, TP2: p<.001) were connected with lower SPQ scores. The current country of residence in Germany significantly associated with lower scores only in the October sample (TP1: p =.677, TP2: p<.001). In contrast having children was related to lower outcomes only in the May sample. (TP1: p =.001, TP2: p =.573). In the May samples, living in a suburban (p =.035) or rural area (p =.025) compared to a big city were connected with increased SPQ scores, while in the October sample, living in a small town (TP1: p = .001) or rural area (TP1: p < .001) compared to a big city had a decreasing effect on SPQ scores. In addition, there was a trend towards an increased SPQ score when living in a suburb compared to a large city (p = .074). See Table 4.

**Tab. 4.**
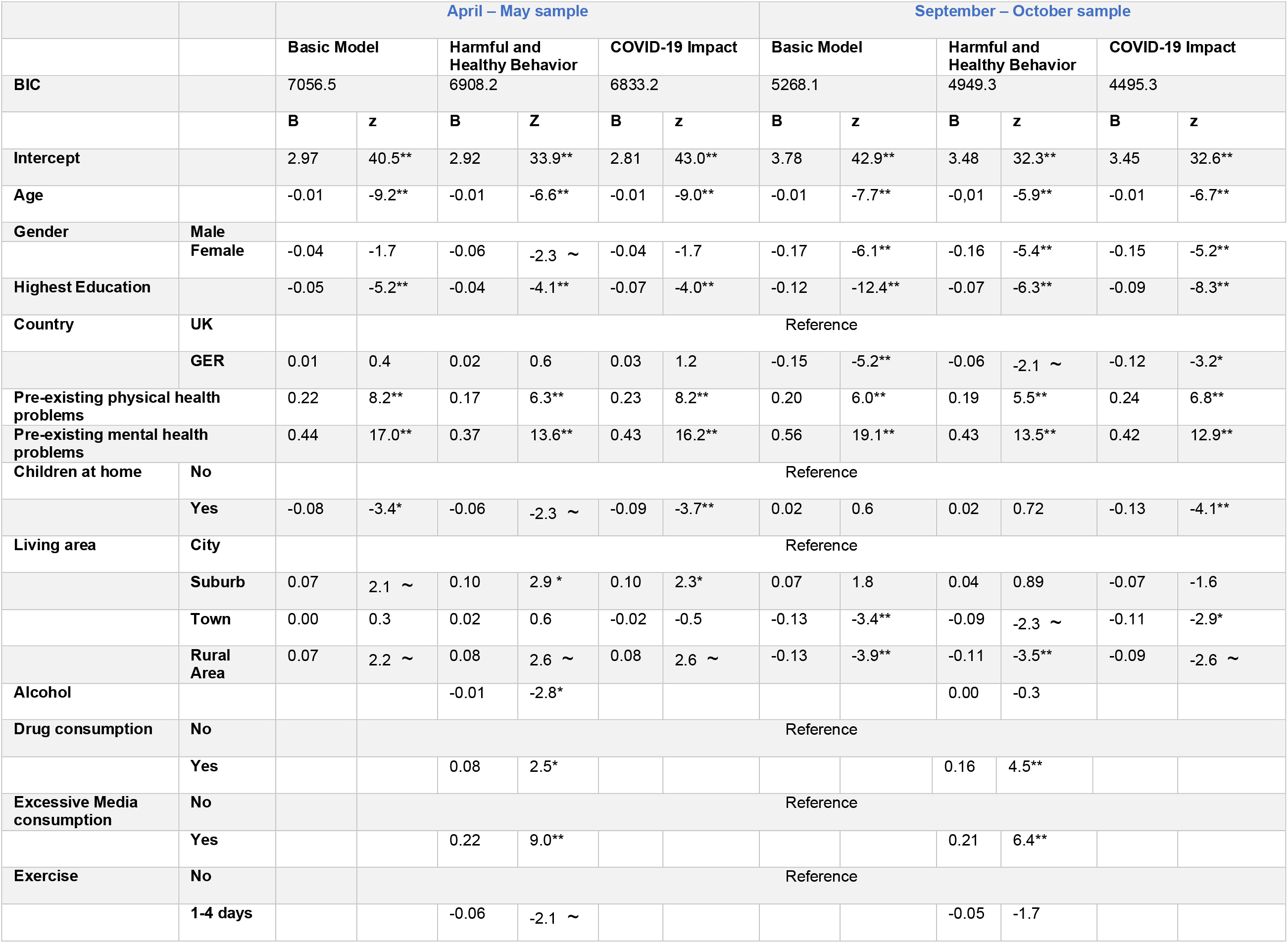
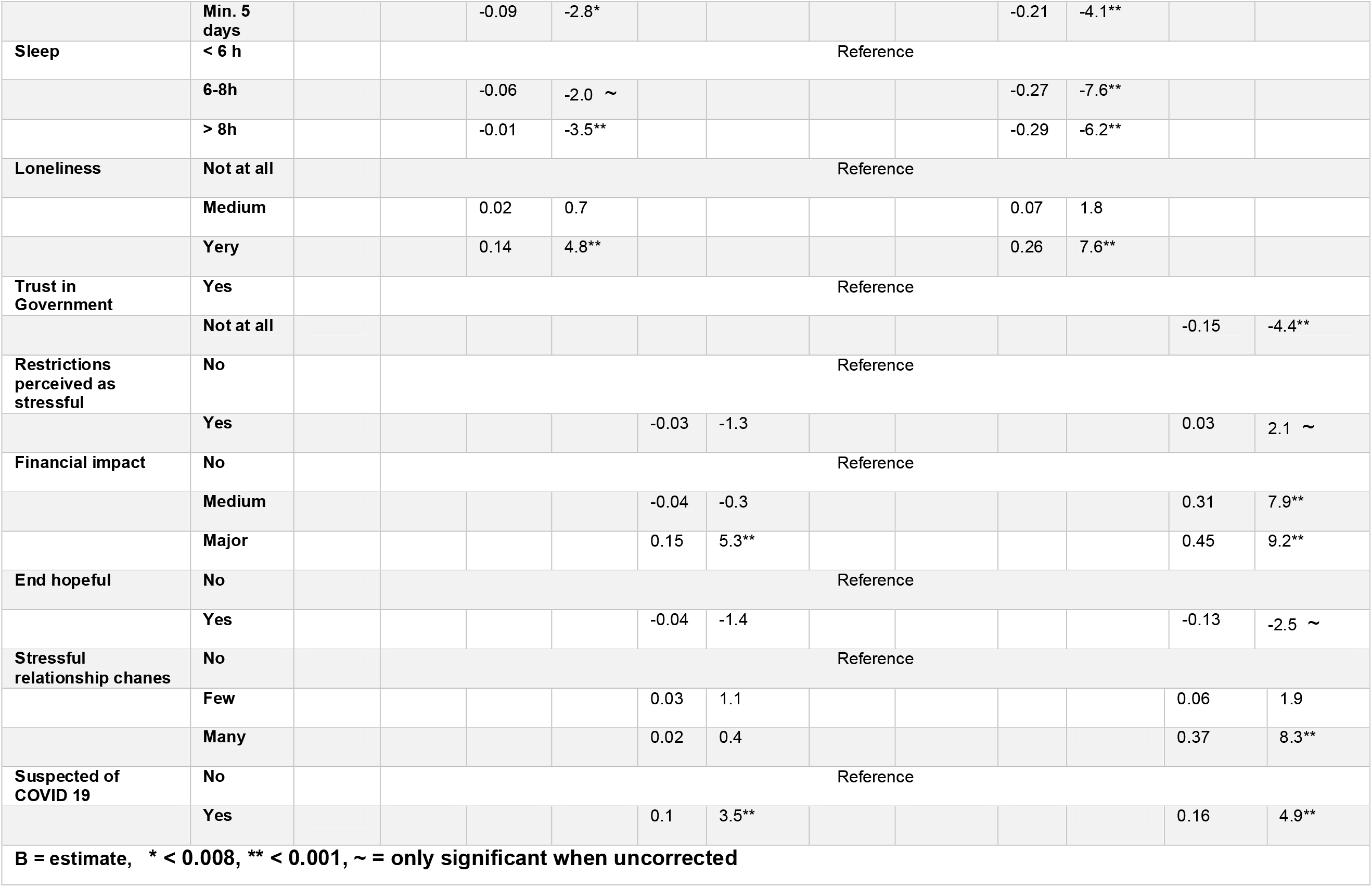
Overview over all three conducted models (Basic Model, Harmful and Healthy Behavior, COVID-19 Impact) for both samples for SPQ scores.

In the two subsequent models, we adjusted for age, gender, country of residence, place of residence, having pre-existing physical and mental health problems as possible confounding variables. All three models have been calculated for the fully-overlapping sample, see supplementary materials, which are comparable to the full sample.

#### 3.4.2 Effects of harmful and healthy behaviour SPQ scores (Harmful and Healthy Behaviour Model)

After adjusting for possibly confounding variables drug consumption (May samples: p<.001, October samples: p<.001) as well as excessive media use (May samples: p<.001, October samples: p<.001) were associated with higher SPQ scores in both samples, whereas alcohol consumption (p =.004) and medium physical exercise (p=.036) were connected with lower scores only in the first sample. Physical Exercise at least five times per week had a negative relationship in both, the May and October samples (May samples: p=.002, October samples: p<.001). Sleeping between six to eight hours (May samples: p=.001, October samples: p<.001) and more than eight hours (May samples: p<.001, October samples: p <.001) both were associated with lower SPQ-scores compared to sleeping less than six hours. See Table 4 and supplementary materials for results of the fully overlapping sample.

#### 3.4.3 Effects of the COVID-19 pandemic on SPQ scores (COVID-19 Impact Model)

After adjusting for confounds, there was a positive relationship of mistrust in Government of leading the country successfully out of the crisis in the October sample (p <.001). Perceiving the restrictions as stressful (p =.043) and being hopeful for a soon end (p =.022) only had negative effects on SPQ scores in the October sample. Medium financial impact of the CRISIS only had a significant association in the second survey (p<.001), whereas major financial were positively correlated with SPQ scores in both, the May and October samples (May: p <.001, October: p <.001). Very stressful relationship changes had only a significant link with the outcome in the second survey (p <.000). The suspicion or diagnosis of being infected with COVID 19 was associated with an increase of SPQ scores in both, the May and October samples (May samples: p <.001, October samples: p =.001). See Table 4 and supplementary materials for results of the fully overlapping sample.

## 4. Discussion

The current study investigated the association of the COVID-19 pandemic with mental health generally and schizoptypy specifically in different, partially overlapping general population samples from the UK and Germany assessed at two time points – the first during widespread societal restrictions aimed at curbing the spread of the virus (April/May 2020), and the second at a time when the majority of these restrictions had been lifted (September/October 2020). Although we are measuring two timepoints, it is not a longitudinal approach, as the samples at both timepoint are only partially overlapping. We are therefore assessing the impact of the pandemic on independent samples from the German and British general population collected in May 2020 and October 2020. The sample from May and October are independent, but highly comparable in terms of distribution of age, and gender. The subjective impact on mental health was quantified using an online survey including questions on the impact on life circumstances, as well as two psychological questionnaires, the Symptom Check List (SCL-27) assessing general psychological symptoms, including depressive symptoms, and symptoms of anxiety, and the Schizotypal Personality Questionnaire (SPQ), assessing schizotypy traits. Furthermore, we assessed the social and economic impact of the pandemic.

We found that the general psychological symptoms (mainly depressive and anxiety symptoms) measured using the Global Symptom Index (GSI, main measure of SCL-27) was significantly lower in the May compared to the October sample in both countries. We were able to confirm these results when running the same analyses in a small sample comparing only those individuals who had taken part at the survey at both timepoints. While during the first timepoint 25-68% of responders were laying above the clinical cut-off for further psychological investigation based on the sub-scores of the SCL-27, at the second timepoint only 12-40% of responders were above clinical threshold. In a normative sample the 10-15% of a cohort reach or exceed the clinical cut-off (Hardt et al., 2004). Schizotypy, however, was higher in the October compared to the May sample, by 4 points in UK responders, 13.6 to 17.4, and by 1 point in German responders, from 12.3 to 13.2. Furthermore, we investigated the subdimensions of the SPQ. The current literature on the SPQ does not provide a consensus on an optimal dimensional structure of the SPQ. In addition to the original nine-factor structure (Raine, 1991), studies have identified a three-factor structure (Raine et al., 1994; Badoud et al., 2011), four-factor structure (Stefanis et al., 2004; Oezgen and Grant, 2018), a bifactor structure (Preti et al., 2015) as well as a six-factor structure (Davies, 2017).This inconsistency is problematic and may arise because the items of the SPQ introduced measurement error. For completion we investigated the six-, nine-, three- and four-factor models. All models show distinct differences between the two countries, especially in overlapping domains such as social anxiety and the interpersonal scores, where the UK scores significantly higher than Germany. Interestingly, this analysis shows that while all scores for the subdimensions for the UK are stable or higher in October compared to May, there is more variation in the score patterns in the German sample.

The results displaying the differences in psychological symptom and schizotypal trait scores confirm our hypotheses. On the one hand, we found that general psychological symptoms (depressive symptoms and symptoms of anxiety) are significantly lower or stay the same comparing samples from October and May, as reported in other studies (Fancourt et al., 2020; Wang et al., 2020). Fancourt and colleagues (2020) report for an only-UK cohort that symptoms depression and anxiety stabilised with the introduction of lockdown easing measures from July 2020, whereas we detect a clear decline in symptoms strength. This might be explained by timepoint of data collection, which was conducted in two months after the Fancourt study, in September/October 2020. The ability to have social contacts, to resume one’s profession, to send children to child care, etc might have a direct alleviating effect. This shows the possibility that the measured increase in symptoms of anxiety and depression at the onset of the pandemic also resembles a normative psychological response to an exceptional situation. Investigating the sub-scores of the symptom check list (SCL-27) in our study, we found the strongest decrease in agoraphobic symptoms; in the UK sample, these symptoms decrease by 20% and in the German sample by 10%. This sub-score of the SCL-27, specifically assesses phobic fears of being among others or supressing actions that could create risks for one’s health, like going outside. These behaviours are expected responses during a pandemic, and are therefore likely to reduce when the risk of infection goes down. The only sub-score of the SCL-27 which increased where vegetative symptoms, like dizziness, heart racing, stomach ache, sickness etc. These symptoms strongly relate to psychosomatic symptoms, which have been reported to have increased significantly in front-line workers (Marinaci et al., 2020; Yi et al., 2020).

On the other hand, and as predicted, we found that schizotypy scores remained the same or were higher at the later timepoint. This is highly interesting, considering that already during the first timepoint nearly 10% of the responders indicated a subjective increase of schizotypy. Recent work shows the impact of adverse life events or loneliness on developing psychotic-like experiences (Beards et al., 2013; Chau et al., 2019; Le et al., 2019; Betz et al., 2020). The social and life-changing consequences of this pandemic (i.e., general reduction of social interaction, job insecurity, experiencing health problems) might therefore provide a long-term risk of schizotypyal trait exacerbation in those individuals with high schizotypy scores. Our regression models indicate that indeed loneliness, financial hardship, and drug consumption are predictors for SPQ. The estimates of those three predictors were increased in the October compared to the May sample. In keeping with prior suggestions (Preti et al., 2020), individuals with increased schizotypal traits might show stronger avoidance behaviours, stronger disruption of social contacts, and with that a delayed return to normality, and therefore take longer to reverse the habits established during the first lockdown showing a worsening of schizotypy scores and a delay to return to baseline. However, this hypothesis requires rigorous longitudinal investigations.

In order to identify associations of the impact of the pandemic on psychological symptoms and schizotypy, we ran three sets of regression models separately for the two timepoints. For GSI, we first setup a basic model: During the first survey we identified positive relations between age, being female, living in the UK, reporting lower mental and physical health prior to the pandemic and living in a town compared to a big city as risk factors, showing an in strong positive association. In the October samples, we identified an additional positive association with lower education. These results, except for living in the UK, confirm previous findings (Adamson et al., 2020; Bäuerle et al., 2020; Bu et al., 2020; Fancourt et al., 2020; Smith et al., 2020; Proto and Quintana-Domeque, 2021; Simha et al., 2021). In the harmful and healthy behaviour model adjusting for the significant factors of the basic model, we investigated harmful and healthy behaviour. We identified the same lowering and increasing association with the outcome in both, the May and October samples. Excessive media consumption and drug consumption contributed to an increased GSI, while longer sleep (>6 hours) was negatively associated. Interestingly the effect of drug consumption is twice as high in the second than the May samples. A recent study showed a strong association between newly initiated substance use and increased measures of COVID related fear and worry (Rogers et al., 2020). Those individuals with highest use and fear and worry scores used substances as necessary coping strategies, which might provide an explanation for the increased association between drug use with GSI in the October sample in our study. Regular sleep of more than 6 hours and healthy sleep routines are usually predictive of better mental health (Milojevich and Lukowski, 2016), it is therefore not surprising that this is the same during a pandemic. Furthermore, we found that excessive media consumption predicts GSI, which confirms previous findings (Bendau et al., 2020). In the COVID-19 impact model adjusting for the significant factors of the basic model we investigated social and economic impact of the COVID-19 pandemic. We found that while the restrictions themselves and the change in social contacts posed a strong stressor during the May survey it was mainly the financial impact, the change in social contacts and the increased risk of infections which posed the greatest influence during the second timepoint. Already during the first peak in April, Witteveen and Velthorst (Witteveen and Velthorst, 2020) linked economic hardship to increased levels of depression and anxiety. During the first peak the economic burden might still be compensated for, however, with the continuing pandemic this burden increases and significantly contributes to mental health decline.

We ran similar regression models to detect potential predictors for schizotypy. In the basic model, we identified similar predictors in both, the May and October samples. While higher age, higher education, and being female were associated with lower outcomes in both, the May and October samples, mental and physical health status before the pandemic were positively correlated. The connection of having children with SPQ scores changed with the continuing of the pandemic from being a negative to a positive predictor; being a UK resident also correlated with higher outcome scores in the October sample. Living in towns or rural areas was associated with lower scores compared to big cities. The link of annual income with SPQ scores was only recorded during the second survey, with increasing effects on the outcome. Gender differences and younger age have been associated with schizotypy previously (Bora and Baysan Arabaci, 2009), and urbanicity (Fett et al., 2019) as well as lower socioeconomic status (Loch et al., 2017) are often linked to psychotic-like experiences.

In the harmful and healthy behaviour model we examined whether harmful and healthy behaviours predicted schizotypy. Adjusting for the significant factors of the basic model, we found the same predictor for both timepoints. While excessive media consumption and drug consumption were linked with higher schizotypy, more exercise and sleep above 6 hours showed the opposite relation. Interestingly, however, the association of drug consumption doubled in the October samples and the connection of more exercise tripled in the October samples. The effect of drug use on schizotypy confirms earlier findings reporting that regular cannabis users score higher on schizotypy and psychosis ratings (Nunn et al., 2001). However, it is a critical finding as drug use is also associated with higher conversion from schizotypy to psychosis (Hjorthøj et al., 2018). Therefore, these results are clinically relevant and requires attention in the course of the pandemic. Regular exercise has been identified as an alleviating intervention for early psychosis (Firth et al., 2018), and should be promoted rigorously during a crisis like the current one.

The COVID-19 impact model investigates the relationship of COVID-19 related measures. Here, we see significant worsening comparing the first and the October samples. The association of financial hardship triples, which is independent from annual income. This might show that financial hardship creates a stressor which imposes a risk not only in people with lower socioeconomic status (Loch et al., 2017), but across a wider range of socioeconomic statuses. Furthermore, stress related the change in social contact more than doubled in the October vs the May samples. This might have been expected that with the continuing course of the pandemic, social isolation might increase, and with that, potentially loneliness too. Loneliness significantly interacts with schizotypy, and has been clinically linked to risk-for psychosis (Chau et al., 2019; Le et al., 2019).

In all models we included country of residence as a predictor, which was significant in most of the models for the May samples and in all models of the October samples. In order to fully understand this relationship, we ran the same regression analysis without country of residence as a predictor (see supplementary Table 6 and 7 for results). All main findings remain the same when excluding the country of residence from the models, suggesting that the overall associations, and especially the directionality, is comparable across both countries, but slightly increased in the UK as indicated by the robust analyses of variance. The reason why UK residents might suffer a stronger mental health burden is multifold. The delayed start of implementing restrictions and due to that the higher numbers of infections and death (Balmford et al., 2020), followed by a higher unemployment and greater loss in gross income (Bauer and Weber, 2020; Mayhew and Anand, 2020), but also general differences in the health care system might contribute (Kuhlmann et al., 2009). Independently, however, the effects are highly similar, which might be due to the comparability of the samples, and the higher proportion of well-trained and socioeconomically-secure responders in both samples.

This study has potential limitations. First, we used online data collection methods, therefore, people without or with limited access to computers, or less well-versed using these methods would be excluded from the sample. However, in order to maximally ease the accessibility of the questionnaire we provided an online version with smart-phone compatible formatting. Second, we used a snowball sampling method for both timepoint with partially overlapping responders, therefore, the sample is not fully representable of the general population. Although we contacted all participants who had completed the first timepoint and agreed to be re-contacted (71.3%), only 121 participants (14.7%) took part in both timepoints. The results of the study should therefore be interpreted considering the sample’s demographics. Furthermore, the reader should be aware that this study is not using a longitudinal approach, it is not showing changes within the same sample. It is however comparing two very similar samples at two timepoints within the ongoing pandemic. Third, comparing two countries is problematic as the countries vary on a large number of factors that are not and cannot be accounted for in detail. Therefore, any differences between the countries presented in this study might be linked to baseline differences. However, by specifically asking for a subjective change considering a pre-verses during-pandemic time-point, we minimised this confound. Fourth, we used a self-reporting survey without clinical in-person verifications. Social distancing measures complicate such verification. However, by using a completely voluntary and anonymous format, as well as standardised questionnaires we are minimising potential effects. Fifth, we are presenting simple regression models without testing for interactions. This approach may not present conclusive results, however, it does allow for comparison with other studies following the same approach, and to generate hypotheses for future research rather than definitive inference. Finally, the usual caveat to observational studies applies, that we are noting associations but cannot infer causality.

In conclusion, we were able to show that whereas general psychological symptoms and percentage of responders above clinical cut-off for further psychological investigation were lower in the sample measured at the second timepoint, following the first peak of the pandemic, schizotypy scores were higher in the October survey. We furthermore found that UK responders were suffering from a stronger mental health burden than responders from Germany. The financial burden, drug use, the impact of loneliness, and previous mental and physical health problems predicted schizotypy, and general psychological symptoms most strongly, but were stronger in the October samples for schizotypy compared to general psychological symptoms. The differences in the scores over time requires further attention and investigations, to understand whether the impact on schizotypy increases further, potentially creating a higher risk for developing psychosis.

## Data Availability

Data available upon request.

## Acknowledgement

We thank the participants of the study who contributed their time and further circulated the survey.

## Notes

Funding FK received funding from the European Union’s Horizon 2020 [Grant number 754462].

Conflict of Interest All authors declare no competing financial interests.

### Competing Interest Statement

The authors have declared no competing interest.

### Funding Statement

FK received funding from the European Union s Horizon 2020 [Grant number 754462].

### Author Declarations

Ethical approval was obtained from the Ethical Commission Board of the Technical University Munich (250/20 S). All respondents included in the analyses provided informed consent.

### Summary of Updates

updated description of statistical results - avoiding an implication of a causal relationship

