## Supplements for "Subjective impact of the COVID-19 pandemic on schizotypy and general mental health in Germany and the UK, for independent samples in May and in October 2020"

**Are we back to normal yet? The impact of the COVID-19 pandemic on mental health with a specific focus schizotypal traits in the general population of Germany and the UK, comparing responses from April/May vs. September/October**

**Sarah Daimer^a^, Lorenz Mihatsch^b^, Lisa Ronan^c^, Graham K. Murray^c,d^, Franziska Knolle^a,c,*^**

**^a^** Department of Diagnostic and Interventional Neuroradiology, School of Medicine, Technical

^b^ Institute for Medical Microbiology, Immunology and Hygiene, Technical University of Munich (TUM), Munich, Germany

^c^ Department of Psychiatry, University of Cambridge, Cambridge UK

^d^ Cambridgeshire and Peterborough NHS Foundation Trust, Cambridge, United Kingdom

1. **Analysis of SPQ subdimensions.**

| Supplementary Table. 1. Overview of means and robust ANOVAS of various SPQ subscales according to Davies et al. (2017), Raine (1991), Raine et al. (1994) and Stefanis et al. (2004) | | | | | | | | | |
| --- | --- | --- | --- | --- | --- | --- | --- | --- | --- |
|  |  | **Mean** | | | |  | **Robust ANOVA/ M-estimator** | | |
|  |  | **April - May** | | **Sept - Oct** | |  | **significance** | | |
|  | **Max** | **UK** | **GER** | **UK** | **GER** |  | **C** | **TP** | **Country x TP** |
| Subscales according to Davies et al., 2017 | | | | | | | | | |
| Anomalous Experience & Beliefs | 18 | 1.74 | 2.41 | 2.19 | 2.10 |  | .053 . | .179 | .104 |
| Social Anhedonia | 15 | 2.97 | 2.70 | 3.78 | 2.95 |  | .638 | .464 | .780 |
| Paranoid Ideation | 11 | 1.65 | 1.89 | 1.83 | 1.83 |  | .001 ** | .267 | .062 . |
| Social Anxiety | 9 | 3.62 | 2.31 | 3.94 | 2.73 |  | .000 *** | .087 . | .342 |
| Eccentricity | 8 | 1.52 | 1.25 | 2.00 | 1.16 |  | .077 . | .057 . | .027 * |
| Disorganised Speech | 7 | 2.15 | 1.69 | 2.44 | 1.69 |  | .000 *** | .119 | .114 |
| Factors according to Raine, 1991 | | | | | | | | | |
| No close friends | 9 | 2.33 | 1.80 | 2.64 | 2.08 |  | .008 ** | .508 | .783 |
| Flattened affect | 8 | 1.54 | 1.33 | 1.93 | 1.48 |  | .250 | .270 | .651 |
| Social anxiety | 8 | 3.18 | 2.07 | 3.46 | 2.25 |  | .000 *** | .087 . | .491 |
| Suspiciousness | 8 | 1.34 | 1.36 | 1.71 | 1.81 |  | .456 | .254 | .102 |
| Magical thinking | 7 | 0.47 | 0.85 | 0.69 | 0.73 |  | i | i | i |
| Unusual perceptual experience | 9 | 1.02 | 0.99 | 1.29 | 0.92 |  | .167 | .396 | .115 |
| Ideas of Reference | 9 | 1.09 | 1.58 | 1.25 | 1.44 |  | .000 *** | .327 | .025 * |
| Odd speech | 9 | 2.42 | 1.98 | 2.92 | 2.00 |  | .001 ** | .466 | .227 |
| Odd behaviour | 7 | 1.33 | 1.06 | 1.71 | 0.95 |  | .013 * | .017 * | .017 * |
| Three factors according to Raine et al., 1994, Four factors according to Stefanis et al., 2004 | | | | | | | | | |
| Interpersonal score | 33 | 8.4 | 6.39 | 9,58 | 7,29 |  | .000 *** | .075 . | .918 |
| Cognitive perceputal score | 28 | 3.91 | 4.80 | 4,74 | 4,39 |  | .307 | .885 | .095 . |
| Disorganised score | 16 | 3.75 | 3.03 | 4,56 | 2,94 |  | .001 ** | .125 | .055 . |
| Paranoia | 24 | 5.61 | 4.85 | 6,45 | 5,18 |  | .160 | .129 | .329 |
| C = Country, TP = Timepoint, . p<.10, * p<.05, **p<.01, ***p <.001, I = interaction, GER = Germany, UK = United Kingdom, i = could not be calculated due to insufficient dispersion or change | | | | | | | | | |

2. **Sub-sample analysis**

127 subjects participated at both time points. 27 of them came from UK and 100 from Germany. Therefore, we additionally calculated the values for SPQ and GSI with this sub-sample in order to exclude the possibility that the changes in the values are only due to the different sample sizes. GSI scores also decreased in this sample (W = 8314, p<.001), while there was no change in SPQ scores (W = 7462, p=.092). We did not investigate the development in the two countries separately, due to the low participation of UK responders. See Supplementary figure 1 and 2 for raincloud plots showing data for timepoints and countries separately.

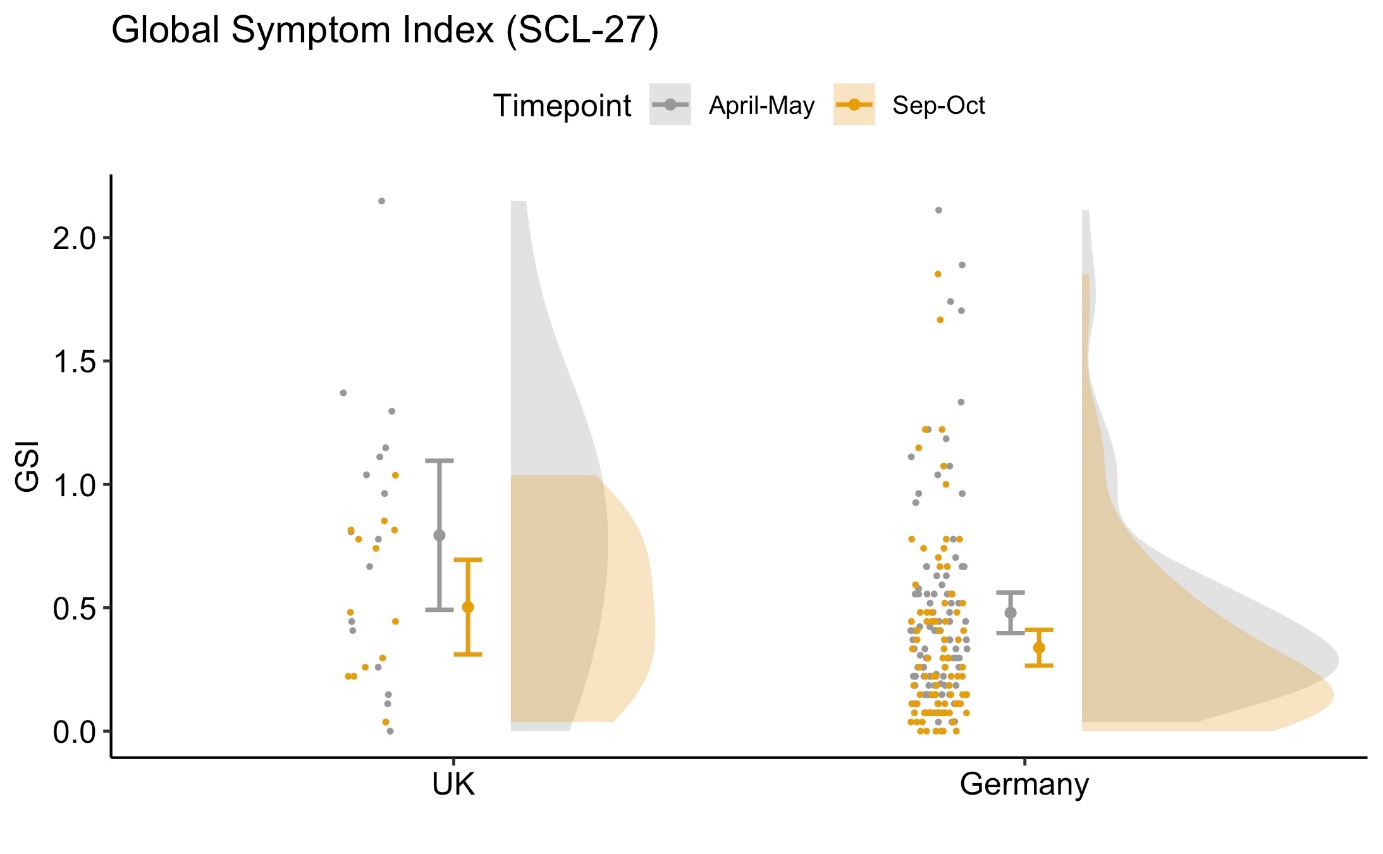
Suppl. Figure 1: Raincloud plot for GSI across country and timepoint for the small, overlapping sample (N=127). Plots show data distribution (the ‘cloud’), jittered raw data (the ‘rain’), mean and standard error.

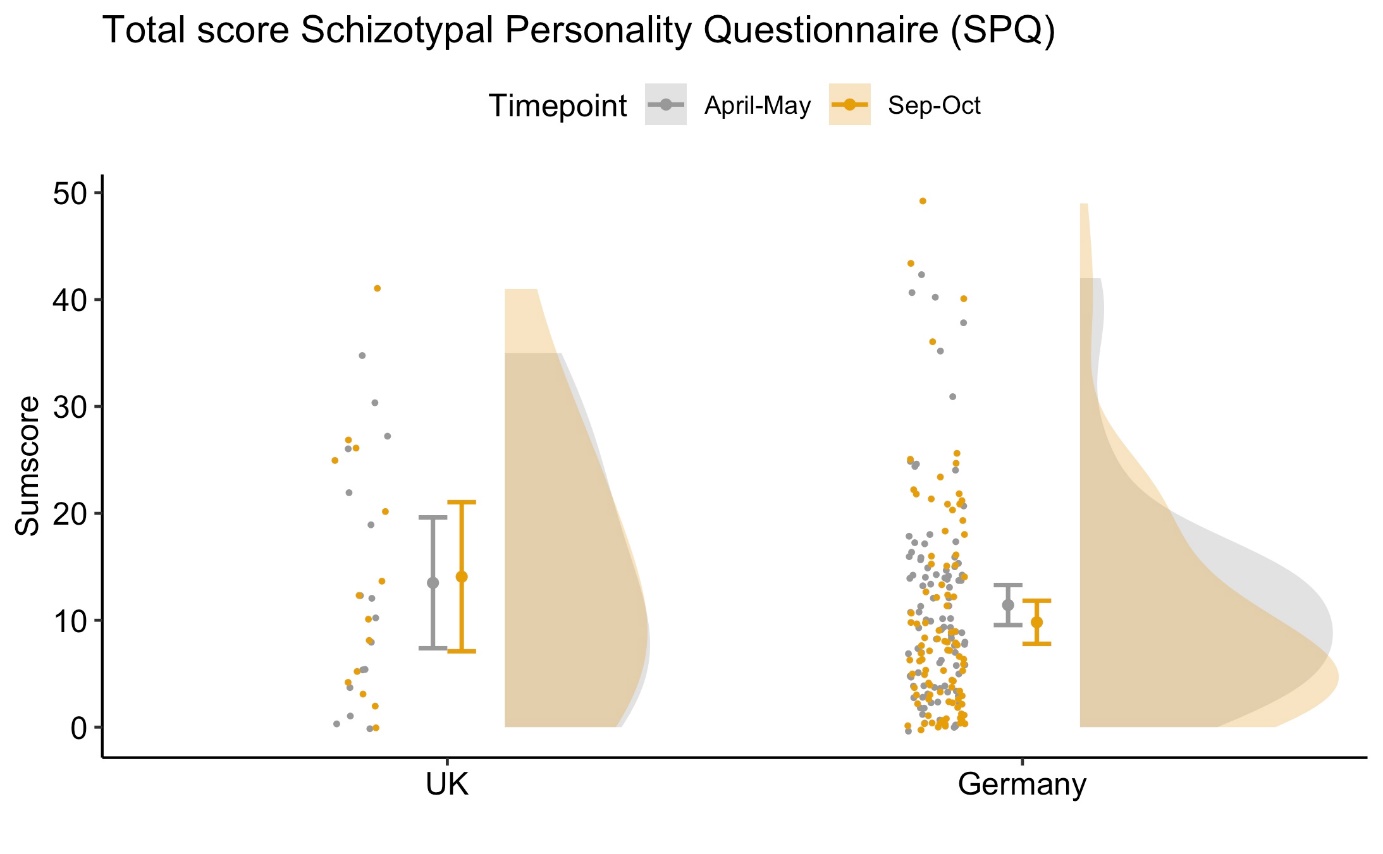

Suppl. Figure 2: Raincloud plot for total SPQ across country and timepoint for the small, overlapping sample (N=127). Plots show data distribution (the ‘cloud’), jittered raw data (the ‘rain’), mean and standard error.

| Suppl. Table 2. Overview over all significant predictor for GSI at the first timepoint | | | | |
| --- | --- | --- | --- | --- |
|  |  | **B** | **t** | **p** |
| (Intercept) |  | 0.58 | 7.43 | .000 *** |
| Age |  | 0.00 | -2.77 | .005 ** |
| Country | **UK** | reference | | |
|  | **GER** | -0.12 | -3.14 | .001 ** |
| Pre-existing mental health problem | | 0.40 | 10.61 | .000 *** |
| Living area | **City** | reference | | |
|  | **Suburb** | 0.13 | 2.60 | .009 ** |
|  | **Town** | 0.13 | 2.94 | .003 ** |
|  | **Rural Area** | 0.10 | 2.48 | .013 * |
|  |  | 0.12 | 3.54 | .000 *** |
| Sleep | **< 6 h** | reference | | |
|  | **6-8 h** | -0.16 | -3.70 | .000 *** |
|  | **> 8h** | -0.17 | -3.35 | .000 *** |
| Restrictions perceived as stressful | | 0.15 | 4.27 | .000 *** |
| Financial impact | **no** |  |  |  |
|  | **medium** | 0.01 | 0.15 | .884 |
|  | **major** | 0.11 | 2.54 | .011 * |
| Stressful Relationship changes | **no** | reference | | |
|  | **medium** | 0.10 | 2.73 | .006 ** |
|  | **major** | 0.20 | 3.70 | .000 *** |
| Suspected of COVID 19 | **no** | reference | | |
|  | **yes** | 0.09 | 2.29 | .022 * |
| Loneliness | **Not** | reference | | |
|  | **medium** | 0.07 | 1.66 | .098 . |
|  | **very** | 0.26 | 5.84 | .000 *** |
| . p<.1, * p<.05, **p<.01, *** p<.001 | | | | |

| Suppl. Table 3. Overview over all significant predictor for GSI at the second timepoint | | | | |
| --- | --- | --- | --- | --- |
|  |  | **B** | **t** | **p** |
| (Intercept) |  | 0.84 | 9.40 | .000 *** |
| Age |  | 0.00 | -4.10 | .000 *** |
| Country | **UK** | reference | | |
|  | **GER** | -0.22 | -5.10 | .000 *** |
| Pre-existing physical health problems | | 0.16 | 2.97 | .003 ** |
| Pre-existing mental health problem | | 0.32 | 6.50 | .000 *** |
| Exercise per week | **0 days** | reference | | |
|  | **1-4 days** | -0.08 | -1.92 | .055 . |
|  | **> 4 days** | -0.14 | -2.22 | .027 * |
| Drug consumption | **Never** | reference | | |
|  | **at least once** | 0.17 | 3.26 | .001 ** |
| Sleep | **< 6h** | reference | | |
|  | **6-8h** | -0.21 | -3.91 | .000 *** |
|  | **>8h** | -0.18 | -2.50 | .012 * |
| Financial impact | **Non** | reference | | |
|  | **medium** | 0.22 | 3.88 | .000 *** |
|  | **major** | 0.55 | 6.88 | .000 *** |
| Stressful relationship changes | **Non** | reference | | |
|  | **few** | 0.14 | 3.16 | .001 ** |
|  | **many** | 0.39 | 6.06 | .000 *** |
| Lonliness | **not** | reference | | |
|  | **medium** | 0.04 | 0.67 | .501 |
|  | **very** | 0.28 | 4.73 | .000 *** |
| . p<.1, * p<.05, **p<.01, *** p<.001 | | | | |

| Tab. 4. Overview over all significant predictor for SPQ at the first timepoint | | | | |
| --- | --- | --- | --- | --- |
|  |  | **B** | **z** | **p** |
| (Intercept) |  | 2.92 | 34.12 | .000 *** |
| Age |  | -0.01 | -6.35 | .000 *** |
| Gender | **Male** | reference | | |
|  | **Female** | -0.06 | -2.54 | .001 ** |
| Highest education | | -0.04 | -4.05 | .000 *** |
| Pre-existing physical health problems | | 0.18 | 6.73 | .000 *** |
| Pre-existing mental health problem | | 0.36 | 13.20 | .000 *** |
| Children | **no** | reference | | |
|  | **yes** | -0.06 | -2.25 | .024 * |
| Living Area | **City** |  |  |  |
|  | **Suburb** | 0.10 | 2.78 | .005 ** |
|  | **Town** | 0.00 | -0,04 | .971 |
|  | **Rural Area** | 0.07 | 2.48 | .013 . |
|  |  | -0.02 | -3.00 | .002 ** |
| Drug Consumption | **never** | reference | | |
|  | **at least once** | 0.08 | 2.63 | .008 ** |
| Excessive Media Use | **no** | reference | | |
|  | **yes** | 0.21 | 8.73 | .000 *** |
| Exercise | **0 days** | reference | | |
|  | **1-4 days** | -0.05 | -1.76 | .078 . |
|  | **>4days** | -0.09 | -2.72 | .006 ** |
| Sleep | **<6h** | reference | | |
|  | **6-8h** | -0.06 | -1.78 | .075 . |
|  | **>8h** | -0.12 | -3.37 | .000 *** |
| Restrictions perceived as stressful | **No** | reference | | |
| Financial impact | **yes** | -0.07 | -2.87 | .004 ** |
|  | **non** | reference | | |
|  | **medium** | -0.06 | -1.68 | .093 . |
|  | **major** | 0.13 | 4.62 | .000 *** |
| Suspected of COVID 19 | **no** | reference | | |
|  | **yes** | 0.07 | 2.50 | .012 * |
| Loneliness | **Not** | reference | | |
|  | **medium** | 0.02 | 0.60 | .550 |
|  | **very** | 0.17 | 5.71 | .000 *** |
| . p<.1, * p<.05, **p<.01, *** p<.001 | | | | |

| Tab. 5. Overview over all significant predictor for SPQ at the second timepoint | | | | |
| --- | --- | --- | --- | --- |
|  |  | **B** | **z** | **p** |
| (Intercept) |  | 3.48 | 31.7 | .000 *** |
| Age |  | -0.01 | -6.17 | .000 *** |
| Gender | **Male** | reference | | |
|  | **Female** | -0.11 | -3.63 | .000 *** |
| Highest education | | -0.06 | -4.85 | .000 *** |
| Pre-existing physical health problems | | 0.24 | 6.92 | .000 *** |
| Pre-existing mental health problem | | 0.35 | 10.77 | .000 *** |
| Children | **no** | reference | | |
|  | **yes** | 0.10 | -3.03 | .002 ** |
| Living Area | **City** | reference | | |
| suburb | **Suburb** | -0.14 | -3.09 | .002 ** |
|  | **Town** | -0.06 | -1.62 | .105 |
|  | **Rural Area** | -0.11 | -3.25 | .001 ** |
| Exercise | **0 days** | reference | | |
|  | **1-4 days** | -0.08 | -2.42 | .015 * |
|  | **>4days** | -0.27 | -4.98 | .000 *** |
| Drug consumption | **Never** | reference | | |
|  | **at least once** | 0.15 | 4.14 | .000 *** |
| Excessive Media consumption | **no** | reference | | |
|  | **yes** | 0.18 | 5.50 | .000 *** |
| Sleep | **<6h** |  |  |  |
|  | **6-8h** | -0.33 | -9.05 | .000 *** |
|  | **>8h** | -0.38 | -7.51 | .000 *** |
| Trust in Government | **yes** | reference | | |
|  | **no** | -0.14 | -4.88 | .000 *** |
| Financial Impact | **non** | reference | | |
|  | **medium** | 0.35 | 8.84 | .000 *** |
|  | **major** | 0.42 | 8.69 | .000 *** |
| Hopeful for soon end | **yes** | reference | | |
|  | **no** | -0.15 | -2.73 | .006 ** |
| Suspected of COVID 19 | **no** | reference | | |
|  | **yes** | 0.17 | 5.12 | .000 *** |
| Stressful relationship changes | **non** | reference | | |
|  | **few** | 0.01 | 0.25 | .806 |
|  | **many** | 0.32 | 7.89 | .000 *** |
| . p<.1, * p<.05, **p<.01, *** p<.001 | | | | |

3. **Correlations**

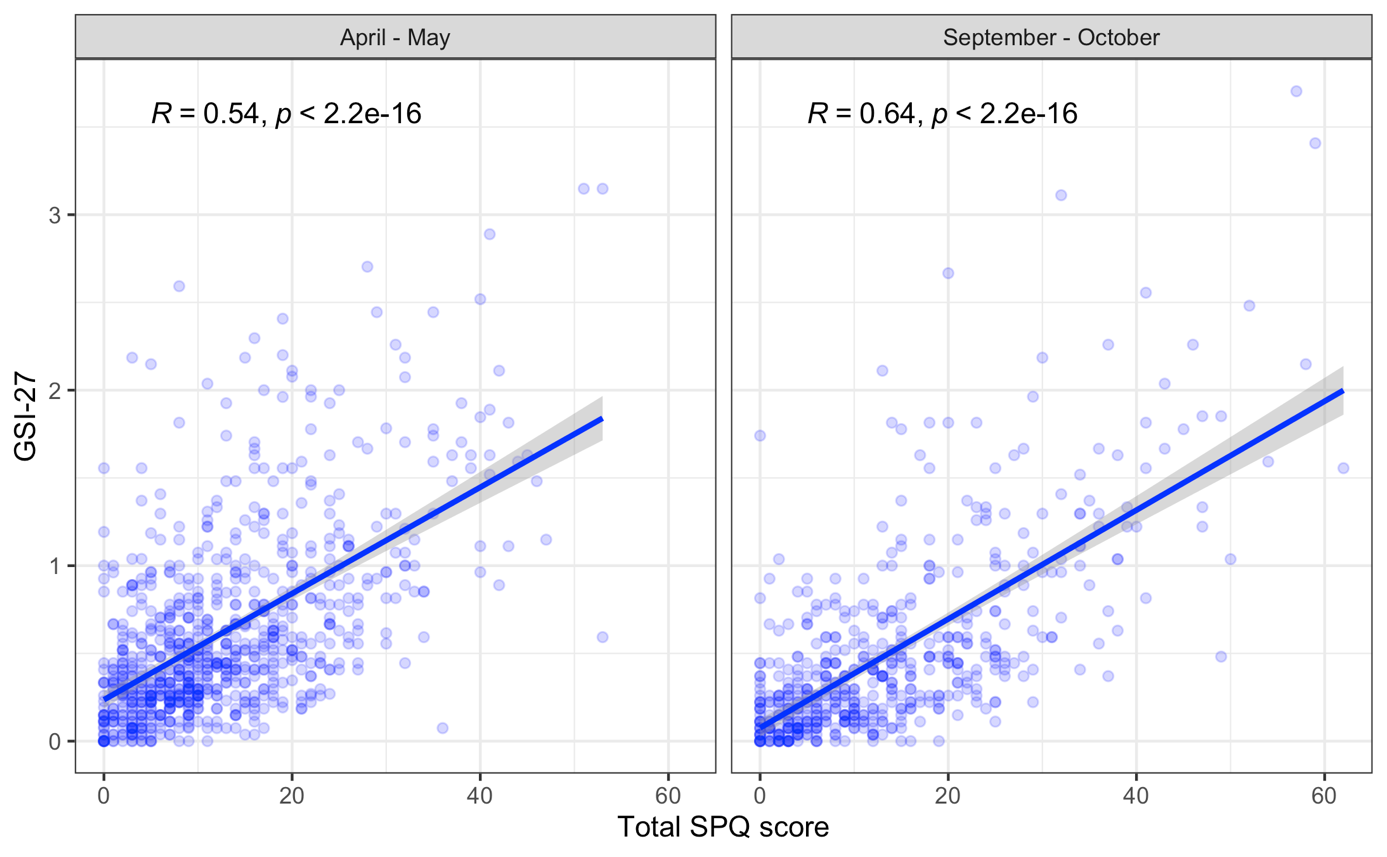

Suppl. Figure 1. Scatterplot for SPQ and GSI-scores separated for both timepoints.

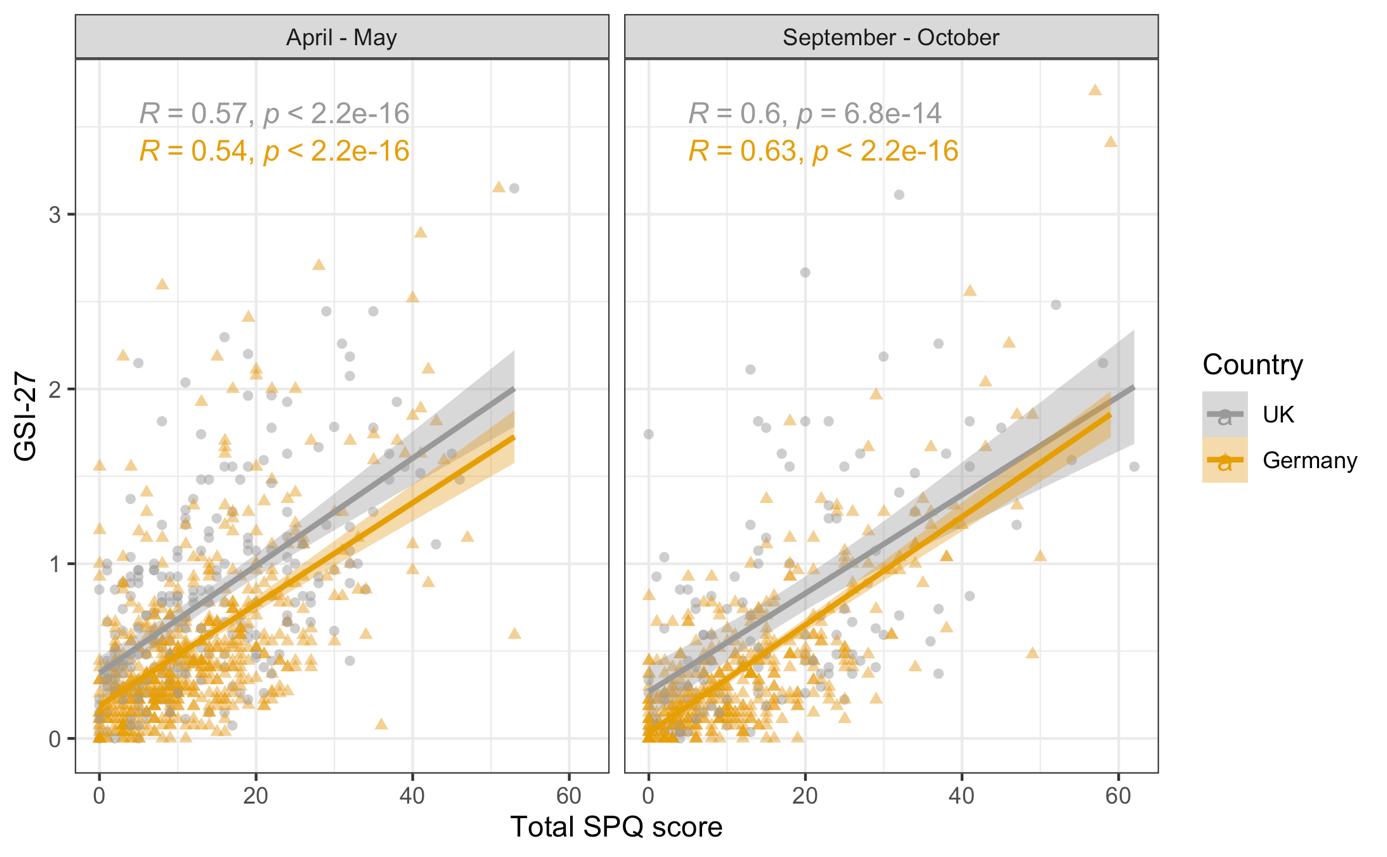

Suppl. Figure 2. Scatterplot for SPQ and GSI-scores separated for both timepoints and Country of residence.

4. **Regression models without country as predictor**

| Suppl. Table 6. Basic Models without Country of residence at both timepoints for GSI scores | | | | | | | | |
| --- | --- | --- | --- | --- | --- | --- | --- | --- |
|  | | **Timepoint 1 (April – May)** | | | **Timepoint 2 (Sept – Oct)** | | | |
| BIC |  | 875.9 | | | 653.6 | | | |
|  | | **B** | **t** | **p** | **B** | **t** | **p** | |
| Intercept |  | 0.57 | 5.01 | <.001 *** | 0.91 | 5.73 | | <.001 *** |
| Age |  | -0.01 | -4.53 | <.001 *** | -0.005 | -3.43 | | .001 *** |
| Gender | **Male** | **Reference** | | | | | | |
|  | **Female** | 0.08 | 2.09 | .037 * | 0.01 | 0.18 | | .855 |
| Highest education |  | 0.00 | 0.31 | .759 | -0.05 | -2.83 | | .005 ** |
| Pre-existing physical health problems | **no** | **Reference** | | | | | | |
|  | **yes** | 0.14 | 2.95 | .003 ** | 0.21 | 3.17 | | .002 ** |
| Pre-existing Mental health problems | **no** | **Reference** | | | | | | |
|  | **yes** | 0.52 | 11.76 | <.001 *** | 0.49 | 8.26 | | <.001 *** |
| Children at home | **no** | **Reference** | | | | | | |
|  | **yes** | -0.06 | -1.49 | .137 | 0.03 | 0.52 | | .606 |
| Living Area | **City** | **Reference** | | | | | | |
|  | **Suburb** | 0.07 | 1.30 | .194 | 0.08 | 1.12 | | .263 |
|  | **Town** | 0.15 | 3.25 | .001 ** | 0.02 | 0.35 | | .728 |
|  | **Rural Area** | 0.08 | 1.81 | .070 . | 0.00 | 0.06 | | .952 |
| B = estimate, . p <.100, * p <.050, ** p <.010, *** p <.001 | | | | | | | | |

| Suppl. Table 7. Basic Models without Country of residence at both timepoints for SPQ scores | | | | | | | |
| --- | --- | --- | --- | --- | --- | --- | --- |
|  | | **Timepoint 1 (April – May)** | | | **Timepoint 2 (Sept – Oct)** | | |
| BIC |  | 7050.2 | | | 5288.4 | | |
|  | | **B** | **z** | **p** | **B** | **z** | **p** |
| Intercept |  | 2.98 | 42.50 | <.001 *** | 3.65 | 43.29 | <.001 *** |
| Age |  | -0.01 | -9.34 | <.001 *** | -0.01 | -7.87 | <.001 *** |
| Gender | **Male** | **Reference** | | | | | |
|  | **Female** | -0.04 | -1.71 | .088 . | -0.18 | -6.33 | <.001 *** |
| Highest education |  | -0.05 | -5.25 | <.001 *** | -0.12 | -12.18 | <.001 *** |
| Pre-existing physical health problems | **no** | **Reference** | | | | | |
|  | **yes** | 0.22 | 8.21 | <.001 *** | 0.22 | 6.58 | <.001 *** |
| Pre-existing Mental health problems | **no** | **Reference** | | | | | |
|  | **yes** | 0.44 | 17.21 | <.001 *** | 0.59 | 20.24 | <.001 *** |
| Children at home | **no** | **Reference** | | | | | |
|  | **yes** | -0.08 | -3.36 | .001 *** | 0.00 | 0.09 | .930 |
| Living Area | **City** | **Reference** | | | | | |
|  | **Suburb** | 0.07 | 2.08 | .037 * | 0.09 | 2.31 | .021 * |
|  | **Town** | 0.00 | 0.15 | .880 | -0.10 | -2.64 | .008 ** |
|  | **Rural Area** | 0.06 | 2.23 | .026 * | -0.10 | -3.15 | .002 ** |
| B = estimate, . p <.100, * p <.050, ** p <.010, *** p <.001 | | | | | | | |
